# Assessment of Androgen Receptor splice variant-7 as a biomarker of clinical response in castration-sensitive prostate cancer

**DOI:** 10.1101/2022.03.10.22271799

**Authors:** Adam G. Sowalsky, Ines Figueiredo, Rosina T. Lis, Ilsa Coleman, Bora Gurel, Denisa Bogdan, Wei Yuan, Joshua W. Russo, John R. Bright, Nichelle C. Whitlock, Shana Y. Trostel, Anson T. Ku, Radhika A. Patel, Lawrence D. True, Jonathan Welti, Juan M. Jimenez-Vacas, Daniel Nava Rodrigues, Ruth Riisnaes, Antje Neeb, Cynthia T. Sprenger, Amanda Swain, Scott Wilkinson, Fatima Karzai, William L. Dahut, Steven P. Balk, Eva Corey, Peter S. Nelson, Michael C. Haffner, Stephen R. Plymate, Johann S. de Bono, Adam Sharp

## Abstract

**Background:** Therapies targeting the androgen receptor (AR) have improved the outcome for patients with castration-sensitive prostate cancer (CSPC). Expression of the constitutively active AR splice variant-7 (AR-V7) has shown clinical utility as a predictive biomarker of AR-targeted therapy resistance in castration-resistant prostate cancer (CRPC), but its importance as predictive biomarker in CSPC remains understudied.

**Methods:** We explored multiple approaches to quantify AR-V7 mRNA and protein in prostate cancer cell lines and patient-derived xenograft (PDX) models, in both publicly available and independent institutional clinical cohorts, to identify reliable approaches for detecting AR-V7 mRNA and protein, and its association with clinical outcome.

**Results:** In publicly available benign prostate, CSPC and CRPC cohorts, *AR-V7* mRNA was much less abundant when detected using reads across splice boundaries than when considering isoform-specific exonic reads. The RM7 AR-V7 antibody had increased sensitivity and specificity for AR-V7 protein detection by immunohistochemistry (IHC) in CRPC cohorts and identified AR-V7 protein reactivity very rarely in CSPC cohorts, when compared to the EPR15656 AR-V7 antibody. Using multiple CRPC PDX models, we demonstrated that AR-V7 expression was exquisitely sensitive to hormonal manipulation. In CSPC cohorts, AR-V7 protein quantification by either assay did not associate with time to development of castration-resistance or overall survival, and intense neoadjuvant androgen-deprivation therapy did not lead to significant detectable AR-V7 mRNA or staining following treatment, and neither pre- nor post-treatment AR-V7 levels associated with the volume of residual disease after therapy.

**Conclusion:** This study demonstrates that further analytical validation and clinical qualification is required before AR-V7 can be considered for clinical use in CSPC as a predictive biomarker.

## Introduction

Prostate cancer is one of the commonest malignancies in men and is a leading cause of male cancer-related death globally (1). The androgen receptor (AR) remains the major therapeutic target in advanced prostate cancer and AR targeting therapies have improved the outcomes of patients with advanced castration-sensitive prostate cancer (CSPC) and castration-resistant prostate cancer (CRPC) (2, 3). Despite these advances, primary and secondary resistance to therapies targeting the AR signaling axis, which include luteinizing hormone releasing hormone analogues, abiraterone, enzalutamide, darolutamide and apalutamide, is inevitable (3). Taken together, the development of analytically validated and clinically qualified predictive biomarkers that identify those patients who benefit from therapies targeting the AR is an area of urgent unmet clinical need; such tests optimize clinical benefit, and quality of life while reducing the treatment related and financial toxicity associated with these treatments.

Resistance to therapies targeting the AR is, in part, driven by the emergence of *AR* amplification and mutations, which invariably impact clinically available AR inhibitors that function through the AR C-terminal ligand binding domain (LBD) (4-8). In addition, constitutively active AR splice variants, of which AR splice variant-7 (AR-V7) is the most frequently observed, is considered both a potential driver of resistance and a clinically validated biomarker for patient treatment stratification in CRPC (9-14). AR-V7 typically arises from alternative mRNA splicing that leads to loss of exons 4-8, and inclusion of cryptic exon 3 (CE3), with the resultant protein product remaining transcriptionally active through its N-terminal domain, whilst lacking the C-terminal LBD (15, 16). AR-V7 drives the growth of prostate cancer cell lines and prostate cancer patient-derived mouse xenografts (PDX) in the presence of therapies targeting the AR (15-17). Furthermore, retrospective clinical studies have demonstrated that AR-V7 mRNA and protein from tissue biopsies and blood (circulating tumor cells and whole blood) are associated with resistance to therapies targeting the AR (9, 11-14, 18). More importantly, a prospective study demonstrated that AR-V7 mRNA-and protein-positive circulating tumor cells are associated with worse outcome (progression free survival and overall survival) of patients with CRPC receiving AR-targeted therapies, although it is important to acknowledge the challenges and limitations of AR-V7 biomarker development in CRPC (10, 19-24).

As AR targeting therapies continue to demonstrate efficacy earlier in disease course the development of predictive biomarkers remains critical (25-30). Although the role of AR-V7 as a predictive biomarker in advanced CRPC is well studied, its relevance in untreated primary prostate cancer is far less clear (31-36). The purpose of this study was to evaluate the significance of AR-V7 expression in primary CSPC. We interrogated the sensitivity and specificity of different measurement approaches to quantify AR-V7 mRNA and protein in multiple prostate cancer models including cell lines and patient-derived models, translating these approaches to multiple independent CSPC patient cohorts with associated clinical outcomes. Here, we demonstrate that AR-V7 is expressed at low mRNA and protein level in untreated primary CSPC compared to CRPC. Using multiple cell lines, patient-derived models, and independent CSPC patient cohorts, we demonstrate in independent laboratories that AR-V7 protein expression determined by immunohistochemistry (IHC) using the CRPC-validated RM7 AR-V7 antibody shows strong specificity for AR-V7. We further show that baseline levels of AR-V7 detected both by IHC or AR-V7-specific splice junctions in RNA sequencing do not predict clinical outcomes in CSPC treated with AR-targeted therapies in multiple clinical settings. Our analyses confirm that in the context of current measurement approaches, AR-V7 expression in untreated, primary CSPC is extremely low with AR-V7 testing therefore unlikely to play no major role as a predicative biomarker in this setting.

## Results

### *AR-V7* mRNA expression is low in primary prostate cancer when spliced reads are evaluated

*AR-V7* has been suggested to be a clinical biomarker in untreated primary prostate cancer; its precise enumeration is therefore paramount. We assessed methods for detecting *AR-V7* transcriptomically by inspecting spliced and unspliced read pileups using the Integrative Genome Viewer (IGV) in multiple datasets. This allowed us to distinguish intronic whole transcriptome sequencing reads that also aligned to the *AR-V7* isoform-defining cryptic exon 3 (CE3) in the intronic regions with reads between exons 3 (Ex3) and 4 (Ex4) of *AR* versus reads that were truly spliced from exon 3 to CE3 (Figure 1A). When *AR-V7* status was defined by evidence of Ex3-CE3 splicing, reads aligning to CE3 were still observed in *AR-V7* negative cases from the benign prostate genotype-tissue expression cohort (GTEx, *n* = 243), primary prostate adenocarcinoma cohort (TCGA-PRAD, *n* = 499), and the Stand-up 2 Cancer-Prostate Cancer Foundation West Coast Dream Team metastatic castration-resistant prostate cancer cohort (WCDT-mCRPC, *n* = 99) (Figure 1B-D) (37-39). Patterns similar to GTEx were also observed in the 52 cases of normal adjacent tissue profiled in TCGA-PRAD (not shown). As anticipated, Ex3-CE3 spliced reads were most abundant in the WCDT-mCRPC cohort (Figure 2A), although across all four cohorts, CE3-aligned reads were significantly more abundant than Ex3-CE3 spliced reads (Figure 1E-G and Supplementary Figure 1A, p < 0.0001, Mann-Whitney test). This observation was not due to any apparent deficiency in splicing detection, as full-length *AR* (defined by splicing from Ex3-Ex4) was observed in all four cohorts (Figure 2B). When normalized by the total number of reads mapped to the *AR* locus (Figure 2C), the WCDT-mCRPC cohort maintained the highest abundance of *AR-V7* spliced reads (Figure 2D). Stratified by *AR-V7* status, only the WCDT-mCRPC cohort demonstrated significant association between CE3-aligned reads and Ex3-CE3 spliced reads (Figure 1H-J and Supplementary Figure 1B, p = 0.001, Fisher’s exact test). We next applied a 59-gene AR-V7 signature score that we have previously shown to be associated with AR-V7 abundance across independent CRPC cohorts (14). Having derived this score from independent CRPC cohorts, we applied it to the WCDT-mCRPC cohort, and demonstrated that *AR-V7* positive patients had significantly (p < 0.001, Welch’s t test) higher AR-V7 signature scores than *AR-V7* negative patients (Figure 2E). Taken together, these important differences in transcriptomic quantification of *AR-V7* abundance impact biomarker detection and suggest that the abundance of reads spanning the Ex3-CE3 splice junction is more accurate than CE3 read counts alone.

**Figure 1.**
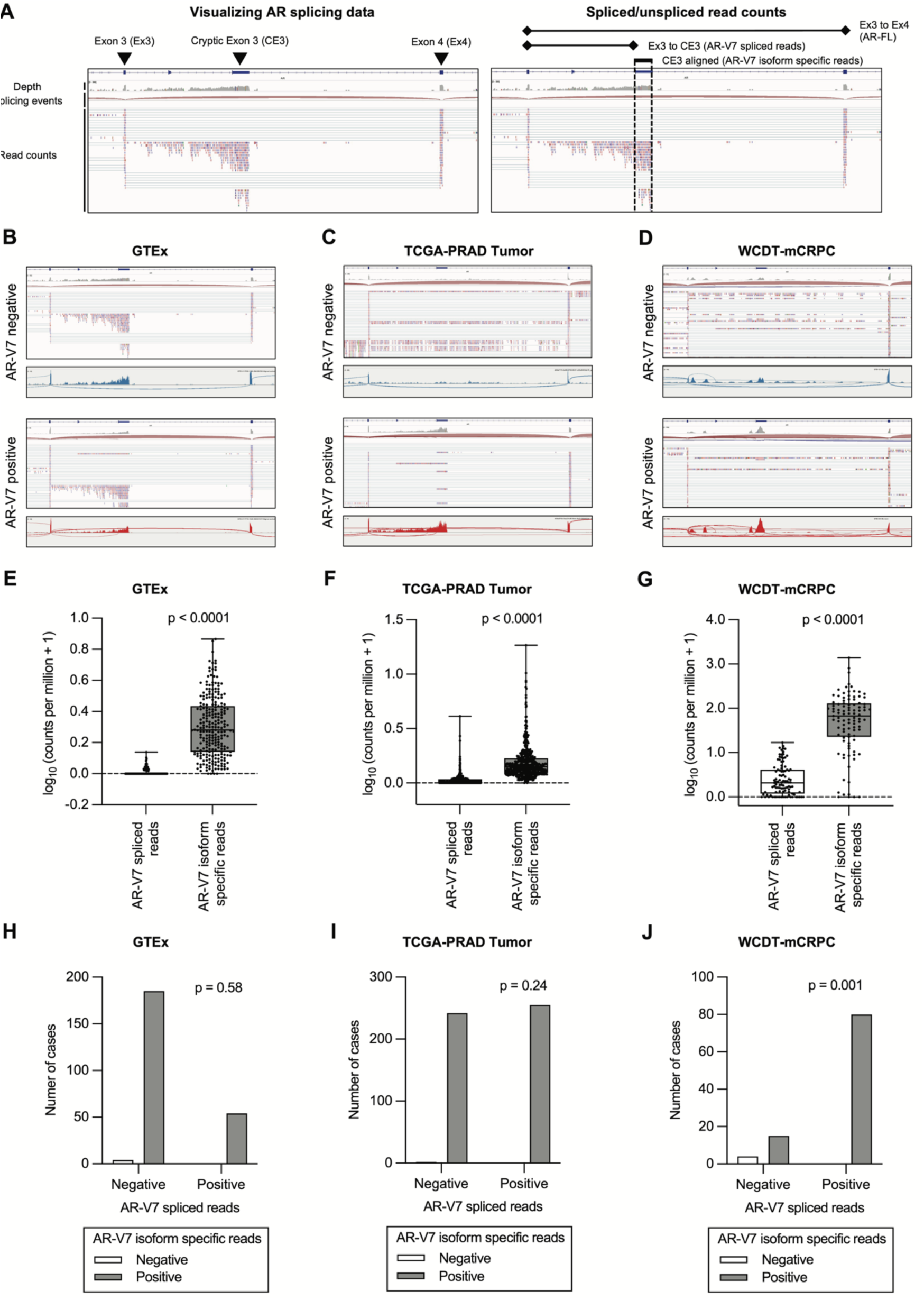
Comparison of AR-V7 quantification in publicly available datasets. **(A)** Screenshot of the Integrative Genomics Viewer (IGV) annotated with information contained in each track with respect to specific regions of the *AR* locus (left) and splice junctions (right). **(B-D)** Screenshots (as in **A**) and sashimi plots for cases from Genotype-Tissue Expression (GTEx, **B**), The Cancer Genome Atlas-prostate adenocarcinoma Tumor (TCGA-PRAD Tumor, **C**) and West Coast Dream Team-metastatic Castration-Resistant Prostate Cancer (WCDT-mCRPC, **D**) cohorts. Representative cases of AR-V7 negative (blue) and AR-V7 positive (red) cases are shown as determined by the presence of splicing between exon 3 and cryptic exon 3 as shown in the red sashimi plots. **(E-G)** For each case in the GTEx (*n* = 243, **E**), TCGA-PRAD Tumor (*n* = 499, **F**), and WCDT-mCRPC (*n* = 99, **G**) cohorts, the number of read counts corresponding to AR-V7 spliced reads (between exon 3 and cryptic exon 3) and AR-V7 isoform specific reads (aligning to cryptic exon 3, dashed vertical lines in **A**) are shown. Spliced reads data are shown as log_10_ (spliced reads per million + 1); isoform specific reads data are shown as log_10_ (read counts per million + 1). Statistical significance between differences were measured by Mann-Whitney tests. Box shows median and interquartile range; bars show minimum and maximum values. (**H-J)** The number of cases showing concordance between the presence of AR-V7 isoform specific reads and the presence of AR-V7 spliced reads is shown for GTEx (**H**), TCGA-PRAD Tumor (**I**), and WCDT-mCRPC (**J**) cohorts. Statistical significance between associations were measured by two-sided Fisher’s exact tests.

**Figure 2.**
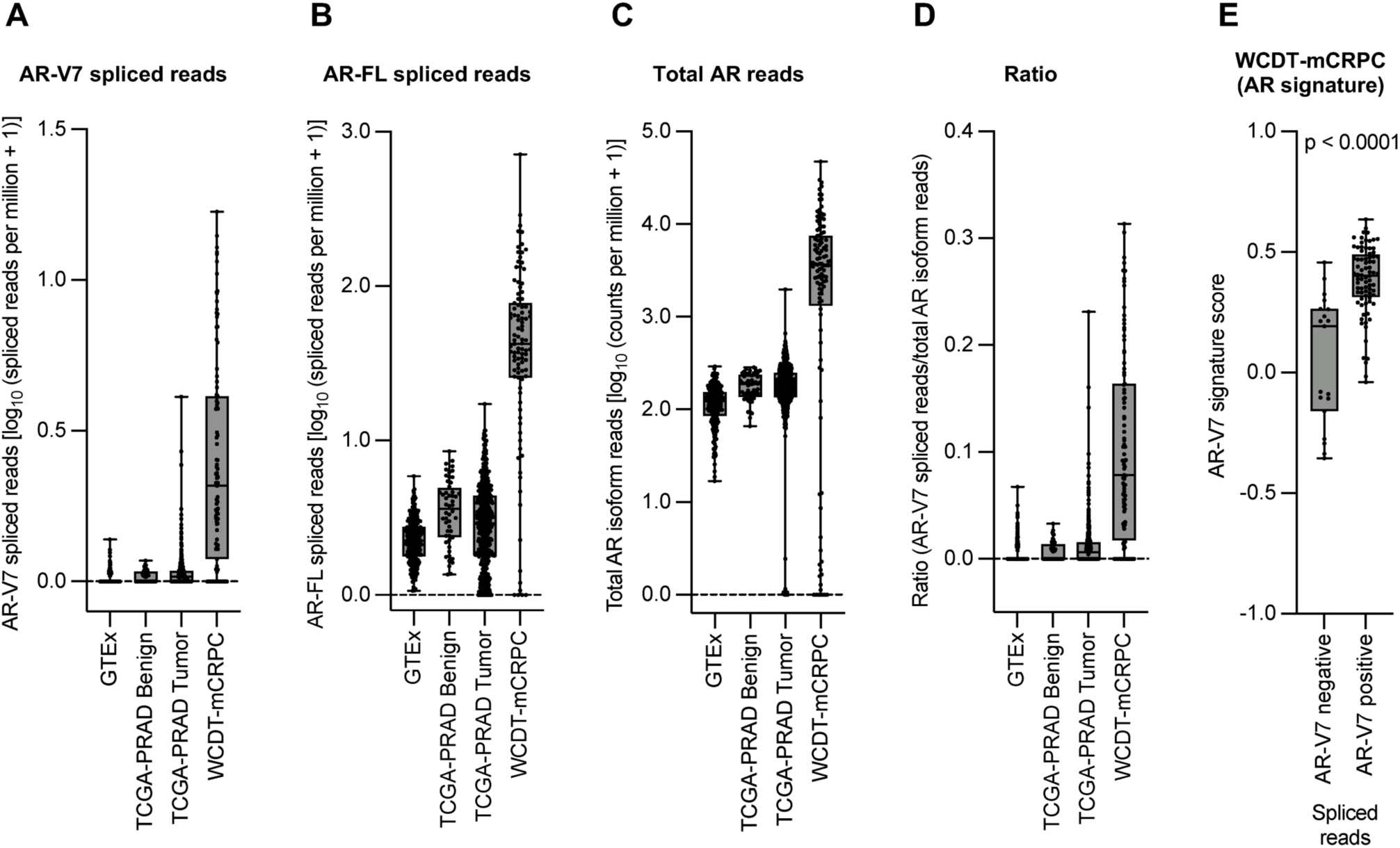
Enumeration of AR isoforms and AR-V7 signature scores in publicly available datasets. **A-D)** The number of reads corresponding to AR-V7 (splicing between exon 3 and cryptic exon 3, **A**), full-length AR (AR-FL, splicing between exon 3 and exon 4, **B**), mapped to the AR locus (**C**), and AR-V7/AR-FL read count ratio (**D**) are shown for the GTEx (*n* = 243), TCGA-PRAD Benign (*n* = 52), TCGA-PRAD Tumor (*n* = 499), and WCDT-mCRPC (*n* = 99) cohorts. Spliced reads data (**A-B**) are shown as log_10_ (spliced reads per million + 1) and isoform specific reads data (**C**) are shown as log_10_ (read counts per million + 1). Box shows median and interquartile range; bars show minimum and maximum values. **(E)** AR-V7 signature score for AR-V7 negative and positive patients determined by AR-V7 spliced reads in WCDT-mCRPC cohort. Box shows median and interquartile range; bars show minimum and maximum values. Statistical significance between differences were measured by the Welch’s t test.

### AR-V7 antibodies demonstrate differences in AR-V7 protein detection in untreated, castration-sensitive, but not castration-resistant prostate cancer

We have previously evaluated two AR-V7 antibodies for immunohistochemistry (IHC), RevMAb RM7 (1 in 500, primary antibody dilution) and abcam EPR15656 (1 in 200, primary antibody dilution) for the detection of AR-V7 protein in prostate cancer tissue biopsies (14, 40, 41). To directly compare these antibodies directly, we utilized castration-sensitive prostate cancer (CSPC) biopsies, with matched castration-resistant prostate cancer (CRPC) biopsies, on which IHC by both antibodies had been performed previously (Institute of Cancer Research/Royal Marsden Hospital matched CSPC and CRPC cohort) (Supplementary Figure 2 and Supplementary Table 1) (14, 40, 41). IHC with RM7 on prostate cancer tissue biopsies demonstrated almost exclusively nuclear staining, while IHC with EPR15656 demonstrated both nuclear and cytoplasmic staining (Figure 3A) (12, 14, 41). Comparing both antibodies, nuclear AR-V7 staining significantly increased (EPR15656, p = 0.009, Mann-Whitney test; RM7, p < 0.0001, Mann-Whitney test) from CSPC (EPR15656, median H-score 35, interquartile range [IQR] 15-90; RM7, H-score 0, IQR 0-0) to CRPC (EPR15656, H-score 90, IQR 42.5-133.8; RM7, H-score 50, IQR 20-120) (Figure 3B). Similarly, using EPR15656, cytoplasmic AR-V7 staining significantly (p = 0.03, Mann-Whitney test) increased from CSPC (H-score 20, IQR 0-70) to CRPC (H-score 70, IQR 10-90) (Supplementary Figure 3). Conversely, and consistent with patterns of exclusively nuclear staining, no significant (p >0.99, Mann-Whitney test) change in cytoplasmic staining between CSPC (H-score 0, IQR 0-0) and CRPC (H-score 0, IQR 0-0) was observed when using the RM7 antibody (Supplementary Figure 3). Staining intensities demonstrated a significant correlation between nuclear AR-V7 staining by EPR15656 and RM7 antibodies in CRPC biopsies (r = 0.60, 0.31-0.79, p = 0.0003, Spearman’s rank) but not CSPC biopsies (r = -0.26, -0.61-0.19, p = 0.23, Spearman’s rank) (Figure 3C), highlighting an important difference in the performance of these antibodies to detect AR-V7 staining in untreated primary prostate cancer.

**Figure 3.**
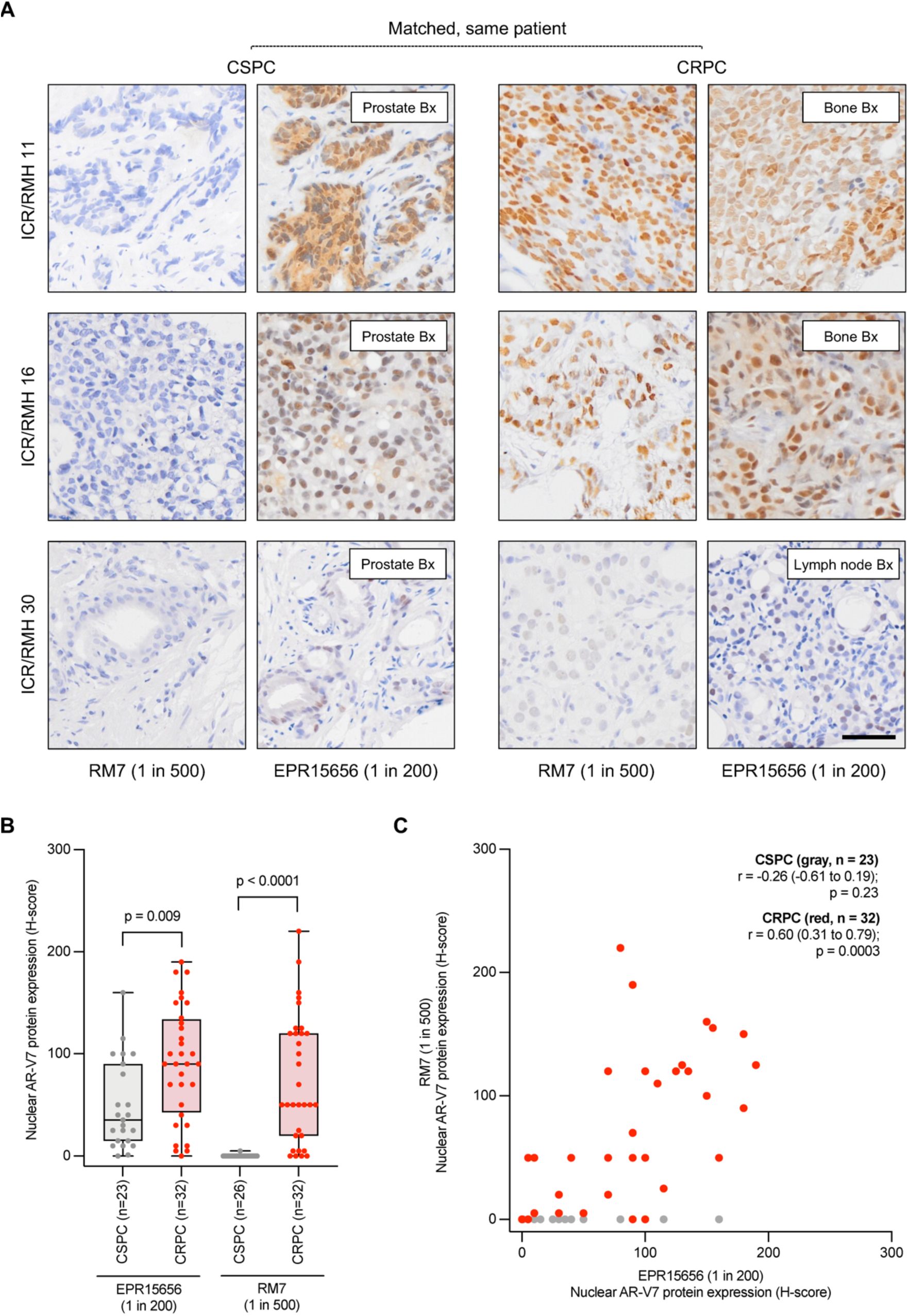
AR-V7 protein quantification by two immunohistochemistry assays in matched, same patient, castration-sensitive and castration-resistant prostate cancer tissue biopsies. **(A)** Representative micrographs of AR-V7 protein detection by immunohistochemistry (IHC) using EPR15656 (abcam, 1 in 200) and RM7 (RevMAb, 1 in 500) antibodies in three patients with matched castration-sensitive prostate cancer (CSPC) and castration-resistant prostate cancer (CRPC) tissue biopsies from the Institute of Cancer Research/Royal Marsden Hospital (ICR/RMH) matched CSPC and CRPC cohort (scale bar: 50 μm). Prostate, lymph node, and bone biopsies (Bx) are shown. **(B)** Nuclear AR-V7 staining (H-score) using EPR15656 (23 CSPC and 32 CRPC) and RM7 (26 CSPC and 32 CRPC) antibodies was determined. Box shows median and interquartile range; bars show minimum and maximum values. Statistical significance between differences were measured by Mann-Whitney tests. **(C)** Nuclear AR-V7 staining (H-score) for EPR15656 and RM7 in the same 23 CSPC (gray) and 32 CRPC (red) biopsies is shown. Statistical significance between correlations were determined by Spearman’s rank.

### Analytical validation of AR-V7 antibodies for immunohistochemistry demonstrates differences in specificity and detection of AR-V7 protein in multiple prostate cancer models

Having demonstrated differences between IHC by RM7 and EPR15656 in prostate cancer tissue biopsies, we next wanted to interrogate further the sensitivity and specificity of both antibodies. We therefore performed IHC on formalin-fixed paraffin-embedded (FFPE) pellets of multiple prostate cancer cell lines. Cell lines previously confirmed to express AR-V7 (LNCaP95, 22Rv1 and VCaP) were positive for AR-V7 by IHC with RM7, while AR-V7 negative cell lines (LNCaP, C42, DU145, PC3 and PNT2) were not positive (Supplementary Figure 4A). While LNCaP95, 22Rv1 and VCaP were also positive for AR-V7 by IHC with EPR15656, the AR-V7 negative cell lines, PC3 and PNT2 demonstrated positivity for AR-V7 by IHC with EPR15656 indicative of non-specific staining (Supplementary Figure 4B). AR protein status was confirmed in all cell lines using both AR N-terminal (NTD) and C-terminal (CTD) antibodies, demonstrating no positivity for AR in PC3 and PNT2 cell lines (Supplementary Figure 4C-D). In addition, western blotting with both RM7 and EPR15656 antibodies demonstrated a strong AR-V7 band at 80kDa in LNCaP95, 22Rv1 and VCaP, but EPR15656 detected a non-specific band in PC3 that was not detected by either AR-NTD or AR-CTD antibodies (Supplementary Figure 5A-D). These data reinforce previous studies that demonstrate both RM7 and EPR15656 recognize AR-V7 protein but EPR15656 may have off-target liabilities that need to be considered (12, 14, 41).

We next performed IHC with both the RM7 and EPR15656 antibodies on VCaP and patient-derived mouse xenograft (PDX) models. Two PDX models were derived from metastatic biopsies of prostate cancer patients with CRPC (Supplementary Figure 6A-B). Once established in intact mice (CP50 and CP89), new castrate (designated C) PDX lines were developed and treated with vehicle or testosterone (20mg/kg OD) for 7 days. RNA sequencing analyses using Ex3-CE3 spliced reads to quantify *AR-V7* demonstrated increased abundance of *AR-V7* mRNA in CP50 and CP89 in response to castration that was reversed by testosterone treatment (Figure 4A-B). IHC by RM7 demonstrated a consistent increase in nuclear AR-V7 staining in tumors grown in intact mice (CP50IV, median H-score 0, range 0-0; CP89IV, H-score 0, range 0-0) compared to castrate mice (CP50CV, H-score 50, range 40-50; CP89CV, H-score 17, range 9-25), and this was suppressed by testosterone treatment (CP50CT, H-score 50, range 40-50; CP89CT, H-score 17, range 9-25) (Figure 4A-B). By contrast, IHC with EPR15656 demonstrated nuclear AR-V7 staining in tumors grown intact mice where *AR-V7* spliced RNA was very low or absent (CP50IV, H-score 135, range 130-135; CP89IV, H-score 90, range 20-95) and there was surprisingly little change when tumors were established in castrate mice (CP50CV, H-score 105, range 45-110; CP89CV, H-score 80, range 50-105), with minimal further change with testosterone treatment (CP50CT, H-score 100, range 95-116; CP89CT, H-score 70, range 27-107) (Figure 4A-B). In addition, cytoplasmic positivity was observed with EPR15656, but not with RM7 (Figure 4A-B). Similar to the PDX’s, staining with RM7 demonstrated an increase in nuclear AR-V7 staining in the VCaP mouse xenograft as it progressed from castration-sensitive (H-score 0, range 0-0) through castration resistance (H-score 105, range 85-125) to abiraterone and enzalutamide resistance (H-score 175, range 140-205) (Figure 4C). However, IHC with EPR15656 also demonstrated nuclear AR-V7 staining at the initial castration-sensitive stage (H-score 60, range 40-165) with only modest increases in intensity with the development of castration resistance (H-score 120, range 60-190) and abiraterone and enzalutamide resistance (H-score 110, range 85-180) (Figure 4C). In addition, as with the PDX models, cytoplasmic positivity was again observed with EPR15656, but not with RM7 (Figure 4C). These data further highlight the differences between IHC with RM7 and EPR15656 in clinically relevant models and demonstrate the dynamic nature of *AR-V7* mRNA and AR-V7 staining that may further complicate its use as a clinical biomarker.

**Figure 4.**
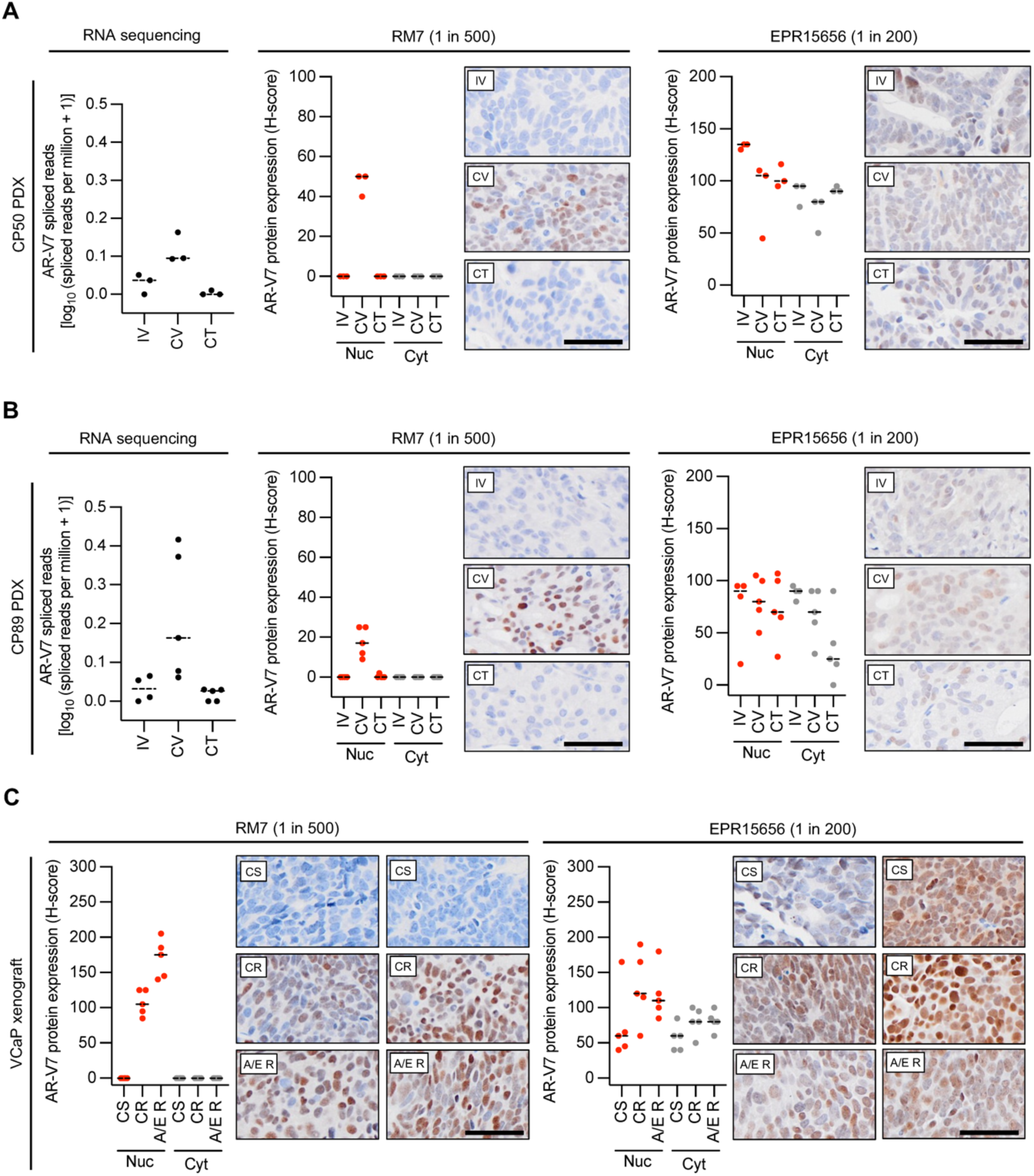
Comparison of AR-V7 immunohistochemistry assays, and AR-V7 spliced reads from RNA analysis, in the CP50 and CP89 prostate cancer patient-derived mouse xenograft, and VCaP mouse xenograft, in response to hormonal manipulation. **(A)** For CP50 prostate cancer patient-derived xenograft (PDX); intact CP50 was treated with vehicle for 7 days (IV, n=3) and its castrate subline CP50C was treated with either vehicle (CV, n=3) or 20mg/kg testosterone daily (CT, n=3) for 7 days and RNA sequencing and immunohistochemistry (IHC) was performed. AR-V7 spliced reads (between exon 3 and cryptic exon 3) were determined from RNA sequencing analysis. Spliced reads data are shown as log_10_ (spliced reads per million + 1). Representative micrographs of AR-V7 protein detection by IHC using EPR15656 (abcam, 1 in 200) and RM7 (RevMAb, 1 in 500) antibodies is shown (scale bar: 50 μm). Nuclear and cytoplasmic AR-V7 staining (H-score) was determined. Line represents median H-score. **(B)** For CP89 prostate cancer patient-derived xenograft (PDX); intact CP89 was treated with vehicle for 7 days (IV, n=4) and its castrate subline CP89C was treated with either vehicle (CV, n=5) or 20mg/kg testosterone daily (CT, n=5) for 7 days and RNA sequencing and IHC was performed. AR-V7 spliced reads (between exon 3 and cryptic exon 3) were determined from RNA sequencing analysis. Spliced reads data are shown as log_10_ (spliced reads per million + 1). Representative micrographs of AR-V7 protein detection by IHC using EPR15656 (abcam, 1 in 200) and RM7 (RevMAb, 1 in 500) antibodies are shown (scale bar: 50 μm). Nuclear and cytoplasmic AR-V7 staining (H-score) was determined. Line represents median H-score. **(C)** For VCaP mouse xenografts; samples were taken from tumors that were castration-sensitive (CS, n=5), as they progressed to castration-resistant (CR, n=5), and as resistance to abiraterone and enzalutamide developed (A/E R, n=5), and IHC was performed. Representative micrographs of AR-V7 protein detection by IHC using EPR15656 (abcam, 1 in 200) and RM7 (RevMAb, 1 in 500) antibodies are shown (scale bar: 50 μm). Nuclear and cytoplasmic AR-V7 staining (H-score) was determined. Line represents median H-score.

We further performed additional IHC with RM7 on the LuCaP series of prostate cancer PDXs for which matched RNA sequencing was also available. Although several castration-resistant (CR) subline models demonstrated an increased abundance of *AR-V7* mRNA, this was not observed in all PDXs (Figure 5A) (42). Independent IHC performed on these models at the Institute of Cancer Research (ICR, 1 in 500, Figure 5B) and University of Washington (UW, 1 in 50) confirmed an increase in nuclear AR-V7 staining in some, but not all, castration-resistant sublines, with more intense staining observed in the UW series. We observed a significant positive association between *AR-V7* mRNA (Ex3-CE3 spliced reads) and both nuclear AR-V7 staining by ICR RM7 IHC (r = 0.64, 0.40-0.79, p < 0.0001, Spearman’s rank) and UW RM7 IHC (r = 0.66, 0.44-0.81, p < 0.0001, Spearman’s rank) (Figure 5D-E). In addition, there was a significant positive association between ICR RM7 IHC and UW RM7 IHC (r = 0.72, 0.53-0.85, p < 0.0001, Spearman’s rank) staining (Figure 5F). Furthermore, LuCaP PDX models that were AR-V7 positive as determined by *AR-V7* spliced reads (p < 0.0001, Mann-Whitney), ICR RM7 IHC assay (p = 0.0001, Mann-Whitney) and UW RM7 IHC (p = 0.0003, Mann-Whitney) had significantly higher AR-V7 gene expression signature scores than AR-V7 negative models (Figure 5A-C). These data demonstrate that quantification of nuclear AR-V7 staining by the RM7 IHC, in two independent laboratories, significantly associates with quantification of *AR-V7* mRNA by Ex3-CE3 spliced reads, and that these measures of AR-V7 quantification associate with an AR-V7 signature score.

**Figure 5.**
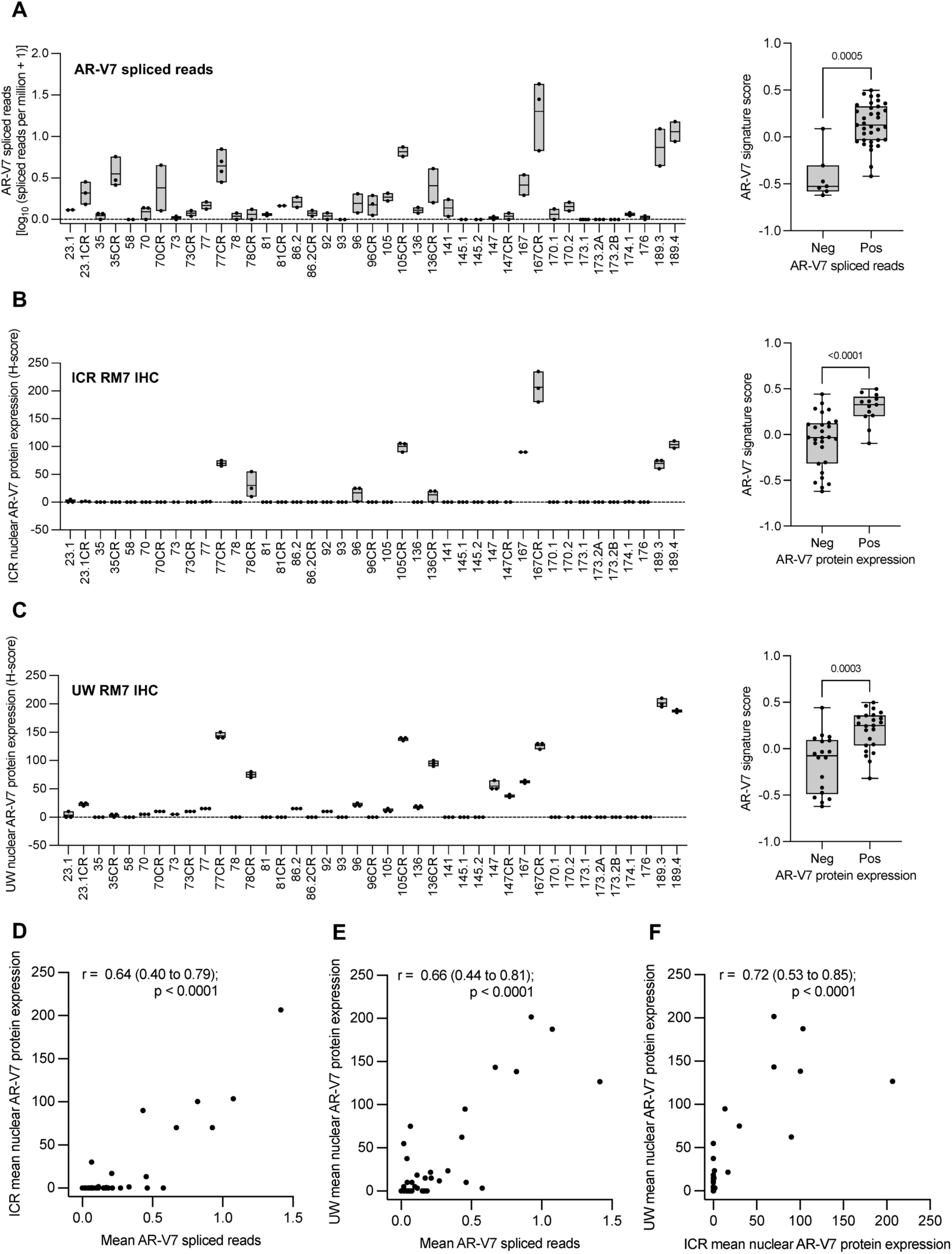
Comparison of AR-V7 immunohistochemistry assays between laboratories, and with AR-V7 spliced reads from RNA analysis, in the LuCaP series of prostate cancer patient-derived xenograft models. **(A)** For each model in the LuCaP series the number of read counts corresponding to AR-V7 spliced reads (between exon 3 and cryptic exon 3) were determined from RNA sequencing analysis. Spliced reads data are shown as log_10_ (spliced reads per million + 1). Box shows mean and bars show minimum and maximum values. AR-V7 signature score shown for AR-V7 negative and positive models determined by AR-V7 spliced reads. Box shows median and interquartile range; bars show minimum and maximum values. Statistical significance between differences were measured by the Welch’s t test. **(B-C)** For each model in the LuCaP series nuclear AR-V7 staining (H-score) was determined by immunohistochemistry (IHC) using RM7 antibody and was performed at the Institute of Cancer Research (ICR, 1 in 500, **B**) and University of Washington (UW, 1 in 50, **C**). Box shows mean and bars show minimum and maximum values. AR-V7 signature score shown for AR-V7 negative and positive models determined by ICR and UW RM7 IHC. Box shows median and interquartile range; bars show minimum and maximum values. Statistical significance between differences were measured by the Welch’s t test. **(D-E)** The association between mean AR-V7 spliced reads and mean nuclear AR-V7 staining determined at ICR (**D**) and UW (**E**) is shown. Statistical significance between correlations were determined by Spearman’s rank. **(F)** The association between mean nuclear AR-V7 staining determined at ICR and UW is shown. Statistical significance between correlations were determined by Spearman’s rank.

Having demonstrated that IHC by RM7 had more intense nuclear AR-V7 positivity at higher concentration, and to increase potential sensitivity to detect AR-V7 protein in primary prostate cancer, we evaluated the sensitivity and specificity of IHC by RM7 and EPR15656 at various primary antibody concentrations in multiple FFPE pellets of prostate cancer cell lines (Supplementary Figure 7A-B). IHC by RM7 remained specific at a 1 in 50 dilution, demonstrating positive and exclusively nuclear staining for AR-V7 protein in AR-V7 positive cell lines (LNCaP95, 22Rv1 and VCaP), and showed no staining in AR-V7 negative cell lines (LNCaP, C42, DU145, PC3 and PNT2) (Supplementary Figure 7A). However, we did identify nucleolar staining that is likely to be non-specific in C42 cells at the 1 in 50 dilution, and thus did not evaluate higher concentrations of RM7 antibody. Although the IHC by EPR15656 demonstrated more intense nuclear AR-V7 positivity in AR-V7 positive cell lines at 1 in 50 dilution that in lower dilutions of this antibody, this intensity was also associated with reduced specificity and off-target positivity in nearly all AR-V7 negative cell lines (Supplementary Figure 7B). These data confirm that EPR15656 has off-target IHC liabilities (12, 14, 41).

Assured of robust sensitivity and specificity of IHC by RM7 at the 1 in 50 dilution, we stained the previously used VCaP and PDX (CP50 and CP89) models at this concentration. At 1 in 50 dilution, IHC by RM7 demonstrated more intense, exclusively nuclear staining, with an increase in nuclear AR-V7 staining from intact state (CP50IV, median H-score 0, range 0-0; CP89IV, H-score 5, range 0-5) to castrate state (CP50CV, H-score 70, range 50-95; CP89CV, H-score 85, range 40-90) that was suppressed by testosterone treatment (CP50CT, H-score 0, range 0-0; CP89CT, H-score 0, range 0-0) (Supplementary Figure 8A-B). We again observed an increase in nuclear AR-V7 staining in the VCaP mouse xenograft as it progressed from castration-sensitive (H-score 0, range 0-0) through castration resistance (H-score 150, range 115-160) to abiraterone and enzalutamide resistance (H-score 180, range 165-205) (Supplementary Figure 8C). Despite the increase in antibody concentration, there was no cytoplasmic staining seen in all models (Supplementary Figure 8A-C). These data further highlight that nuclear AR-V7 staining by RM7 IHC associates with quantification of AR-V7 mRNA by Ex3-CE3 spliced reads.

### Validated AR-V7 immunohistochemistry rarely identifies AR-V7 protein in primary prostate cancer and detection does not associate with clinical outcome

We next explored both IHC with RM7 at 1 in 50 dilution, and EPR15656 at 1 in 200 dilution, in two independent patient cohorts of primary advanced prostate cancer (ICR/RMH primary advanced cohort) and primary localized prostate cancer (UW primary localized cohort) to determine whether AR-V7 protein was identified and associated with clinical response (Supplementary Figure 9, Supplementary Table 2 and Supplementary Table 3). In the ICR/RMH primary advanced cohort comprised of 22 patients who received systemic therapy for primary advanced prostate cancer, only 2 patients (9%) demonstrated nuclear AR-V7 staining, and no patient demonstrated cytoplasmic positivity when tissues were evaluated with RM7 IHC (Figure 6A-B). Nuclear AR-V7 staining at baseline did not associate with time to the development of CRPC (r = -0.16, -0.56-0.29, p = 0.47, Spearman’s rank) or overall survival (r = -0.24, -0.61-0.22, p = 0.29, Spearman’s rank) (Figure 6C-D). By contrast, IHC by EPR15656 identified AR-V7 positivity in primary advanced prostate cancer, with 15 patients (68%) demonstrating nuclear AR-V7 staining, and 6 patients (27%) demonstrating cytoplasmic positivity (Figure 6A-B). Despite increased nuclear AR-V7 staining, there remained no association between nuclear AR-V7 staining at baseline and time to development of CRPC (r = -0.23 -0.60-0.23, p = 0.31, Spearman’s rank) or overall survival (r = -0.15, -0.55-0.30, p = 0.40, Spearman’s rank) (Figure 6C-D).

**Figure 6.**
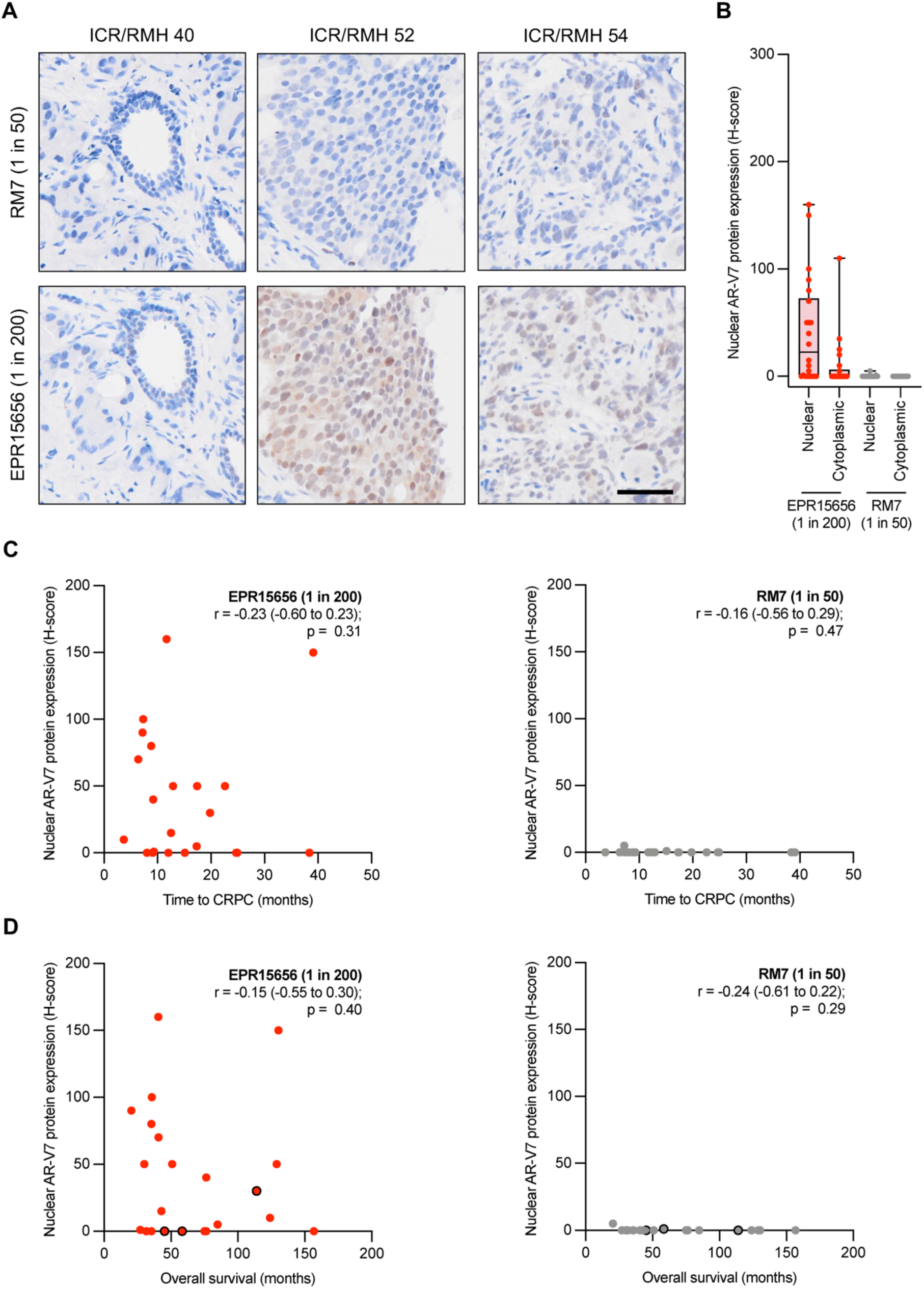
AR-V7 protein quantification by two immunohistochemistry assays in diagnostic biopsies of prostate cancer patients who received systemic therapy alone. **(A)** Representative micrographs of AR-V7 protein detection by immunohistochemistry (IHC) using EPR15656 (abcam, 1 in 200) and RM7 (RevMAb, 1 in 50) antibodies in three diagnostic castration-sensitive prostate cancer (CSPC) prostate biopsies from patients in the Institute of Cancer Research/Royal Marsden Hospital (ICR/RMH) primary advanced cohort (scale bar: 50 μm). **(B)** Nuclear and cytoplasmic AR-V7 staining (H-score) using EPR15656 (abcam, 1 in 200, red circles) and RM7 (RevMAb, 1 in 50, gray circles) antibodies was determined. Box shows median and interquartile range; bars show minimum and maximum values. **(C)** Association between nuclear AR-V7 staining (H-score) using EPR15656 (EPR15656, 1 in 200, red circles) and RM7 (RevMAb, 1 in 50, gray circles) antibodies and time to castration-resistant prostate cancer (CRPC, months) from diagnosis is shown. Statistical significance between correlations were determined by Spearman’s rank. **(D)** Association between nuclear AR-V7 staining (H-score) using EPR15656 (EPR15656, 1 in 200, red circles) and RM7 (RevMAb, 1 in 50, gray circles) antibodies and overall survival (months) from diagnosis is shown. Three patients remain alive at last follow up (black circle outline). Statistical significance between correlations were determined by Spearman’s rank.

This pattern of staining was further validated in the UW primary localized cohort, which consisted of 26 prostatectomies with benign prostate and prostate cancer tissue, and other benign tissue (including kidney, spleen, and salivary gland) (Supplementary Figure 9 and Supplementary Table 3). The prostate cancer tissues from these prostatectomies were included in the 295 samples previously studied with the RM7 antibody (14). We observed no nuclear AR-V7 staining in prostatic adenocarcinoma tumor (H-score 0, IQR 0-0) (14). In addition, further to previous studies, we demonstrate no nuclear AR-V7 positivity in adjacent benign prostate tissue (median H-score 0, IQR 0-0) or other benign tissues (H-score 0, IQR 0-0) (Supplementary Figure 10). Although IHC by EPR15656 demonstrated less nuclear AR-V7 staining in this cohort, reactivity was observed even in benign prostate (median H-score 2, IQR 0-4.5), prostate tumor (H-score 0, IQR 0-1) and other benign tissues (H-score 1, IQR 0-4) (Supplementary Figure 10). These data confirm that IHC by RM7 very rarely detects nuclear AR-V7 staining in both advanced and localized primary prostate cancer tissues at diagnosis, and although positivity is seen with IHC by EPR15656 in primary prostate cancer and other tissue types, this is likely to be related to its off-target liabilities, with EPR15656 staining not associated with clinical outcomes in this patient cohort.

### AR-V7 mRNA quantification by junction-specific reads does not increase in response to neoadjuvant therapy or predict response to therapy in primary prostate cancer

Finally, we assessed whether AR-V7 can predict or mediate resistance to short-course intense androgen-deprivation therapy (ADT) prior to prostatectomy, using tissue samples and RNA sequencing data from a recent clinical trial at the United States National Cancer Institute (NCI) (43). As RNA was extracted from FFPE samples, we first sought to determine whether our ability to detect spliced and total *AR* was affected by these processing steps, using a localized prostate cancer cohort (Supplementary Figure 11A, *n* = 84) of matched benign and tumor blocks subjected to whole transcriptome sequencing. *AR-V7* abundance in both the benign and tumor blocks was comparable to the GTEx and TCGA-PRAD benign and tumor tissues (see Figure 2A) except for a single case that received a short course of bicalutamide prior to prostatectomy (Figure 7A). The NCI neoadjuvant cohort (Supplementary Figure 13B, *n* = 37) similarly showed extremely low levels of Ex3-CE3 *AR-V7* spliced reads in matched pre- and post-treatment specimens (Figure 7B). Moreover, this low abundance of spliced reads observed in biopsy specimens did not track with treatment response as measured by the volume of residual disease after therapy (Figure 7B). This finding was not due to a deficiency in detecting splicing events by RNA sequencing, as both cohorts demonstrated comparable levels of full-length AR (Ex3-Ex4 splicing) to TCGA-PRAD (Supplementary Figure 12).

**Figure 7.**
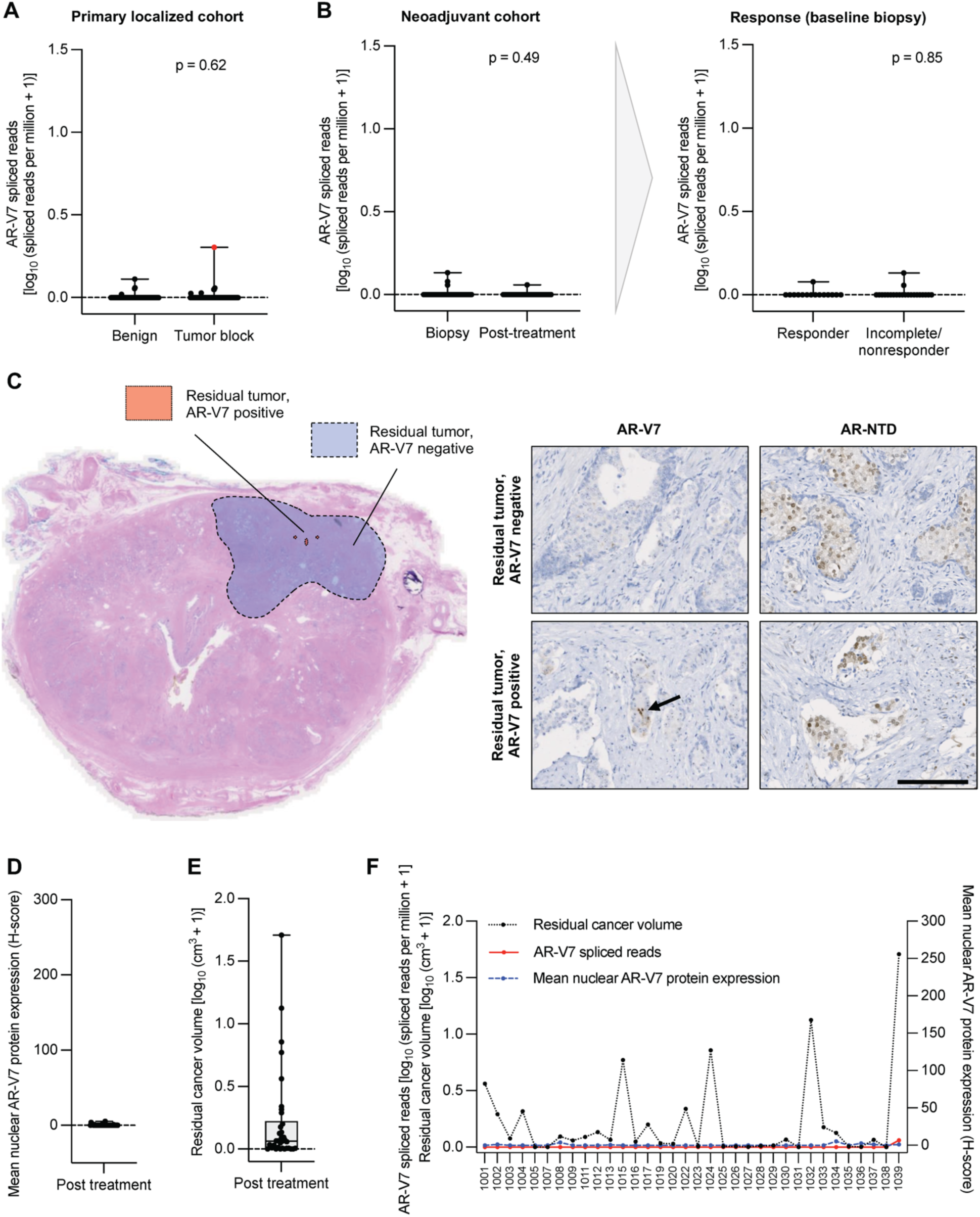
AR-V7 quantification in prostate cancer patients who had primary radical prostatectomy or were treated with neoadjuvant ADT plus enzalutamide for locally advanced disease prior to prostatectomy. **(A)** AR-V7 spliced reads [depicted as log_10_ (spliced reads per million + 1)] from 84 cases treated by radical prostatectomy. The red dot indicates the only case that received short-course neoadjuvant bicalutamide. Box shows median and interquartile range; bars show minimum and maximum values. Statistical significance between differences were measured by the Mann-Whitney test. **(B)** AR-V7 spliced reads [depicted as log_10_ (spliced reads per million + 1)] from 37 cases treated with six months of neoadjuvant androgen-deprivation therapy (ADT) plus enzalutamide prior to radical prostatectomy. Left: Comparison of AR-V7 spliced reads from baseline biopsy and post-treatment. Right: Stratification of baseline biopsies based on pathologic outcome of responder (n=15) or incomplete/nonresponder (n=22). **(C)** Left: Whole slide imaging of a radical prostatectomy specimen from a representative incomplete/nonresponder; region of residual tumor is marked by dotted line. Serial whole-slide section of AR-V7 staining using RM7 (RevMAb, 1 in 100) antibody to identify regions of residual tumor that are AR-V7 positive (small foci marked in red) or AR-V7 negative (larger region marked in blue). Right: Representative micrographs of AR-V7 and AR N-terminal (AR-NTD) immunohistochemistry (IHC) of residual tumor from serial sections (scale bar: 200 μm). **(D)** Distribution of H-scores for AR-V7 IHC from post-treatment specimens with residual tumor (n=34). **(E)** Distribution of residual cancer volumes for each patient receiving six months of neoadjuvant ADT plus enzalutamide prior to radical prostatectomy. Data presented as log_10_ (cm^3^ + 1). **(F)** Overlay of data presented in **B, D** and **E**, displayed by patient with all data available (n=34). AR-V7 spliced read abundance and residual cancer volume plotted on the left Y axis, and mean H-score for nuclear AR-V7 staining plotted on the right Y axis.

In addition to the transcriptomic analysis performed, all post-treatment specimens were evaluated for residual tumor using both full-length AR and RM7 AR-V7 antibodies by IHC. As depicted in Figure 7C, the most common observation was restoration of nuclear AR expression in large volumes of residual tumor (blue shading) with microscopic nests of cells demonstrating rare AR-V7 staining. All post-treatment samples had H-scores less than 10 (Figure 7D). Despite a range of residual tumor volumes after therapy, neither AR-V7 spliced reads nor nuclear AR-V7 staining tracked with residual tumor volume, indicating that AR-V7 levels neither predict nor track with residual tumor volume following neoadjuvant therapy in this clinical study (Figure 7E-F).

## Discussion

The AR remains the major therapeutic target in CRPC (3). AR-targeted therapies have improved overall survival and quality of life of patients with CRPC (2, 3). However, acquired resistance to AR-targeting therapies is common, and is associated with the emergence of AR-V7, a constitutively active AR splice variant at the mRNA and protein levels (9-14, 18, 41, 44). More recently, AR-V7 status as a predictive biomarker has shown clinical utility in guiding treatment with AR-targeted therapy vs. systemic chemotherapy in assays featuring AR-V7 mRNA or protein reactive circulating tumor cells (CTCs) (9-13). However, the development of AR-V7 as a predictive biomarker in CRPC has not been without its challenges. These include: (1) antibody specificity for AR-V7 protein in tissue samples; (2) biomarker absence (AR-V7 negative CTC) being deemed AR-V7 negative; (3) CTC enumeration potentially confounding survival analysis; (4) confirming predictive over prognostic utility; and (5) limited translation to routine clinical care (10-14, 19-24, 41, 44).

The treatment landscape for advanced prostate cancer is rapidly changing with the demonstration that AR-targeting therapies improve the overall survival for patients with advanced CSPC, raising the question of whether AR-V7 may serve as a predictive biomarker earlier in the prostate cancer paradigm (3). However, the evidence supporting AR-V7 as a biomarker in CSPC has been associated with significant challenges that impact its clinical qualification. As shown here and by others, these challenges include the low abundance or absence of AR-V7 in CSPC, CSPC’s high response rate to primary therapy including AR-targeting agents, and the complexity of multi-modal treatment strategies being utilized earlier in the disease course (36). Nonetheless, multiple studies have reported that AR-V7 protein expression was associated with poorer outcome in CSPC (31-35). An important consideration for these studies, and any tissue-based IHC analyses, is the analytical validation to determine the specificity and sensitivity of the antibodies used, which have mainly been RM7 and EPR15656 described in this study (14, 32, 34, 35).

Based on our head-to-head analyses here, and previously, IHC by EPR15656 recognizes AR-V7 protein but also clearly demonstrates non-specificity with cross-reactivity with other undefined proteins in multiple orthogonal validation assays when compared to IHC by RM7 (14). This highlights the importance of careful antibody validation and the use of rigorous controls when new IHC assays are developed and applied (45-47). In addition, although AR-V7 staining by RM7 and EPR15656 antibodies significantly correlated in CRPC tissue biopsies, IHC by EPR15656 commonly showed AR-V7 reactivity in independent tissue cohorts of locally advanced primary prostate cancer, as well as in benign prostate and other benign tissues, compared to IHC by RM7. Furthermore, AR-V7 reactivity by either antibody did not associate with clinical outcome in advanced primary prostate cancer treated with systemic therapy alone. Although is possible that this increased AR-V7 reactivity in primary prostate cancer tissue samples seen with EPR15656 is due to superior sensitivity when compared to RM7, when considering the extensive validation shown here this is highly unlikely to be the case. These data indicate that this detected staining with EPR15656 in CSPC is largely likely to be a consequence of non-specific cross reactivity which may be further compounded by the varying preanalytical variables associated with archival tissues when used (47, 48).

An alternative to IHC is the quantification of segments of *AR-V7* mRNA that are specific to the isoform, using either RNA sequencing or quantitative PCR. Here, utilizing multiple publicly available RNA-seq data sets, we, as have others, demonstrate that *AR-V7* mRNA is commonly identified in benign prostate and primary prostate cancer tissue using isoform specific, CE3 aligned, reads (49). Studies have demonstrated that isoform specific reads significantly associate with, but are much more abundant than, Ex3-CE3 spliced reads (49). Consistent with this, *AR-V7* mRNA was much less frequently observed in benign prostate and primary prostate cancer tissue when Ex3-CE3 spliced reads were examined, with an expected significant increase in *AR-V7* abundance seen in metastatic CRPC. High levels of *AR* expression and the uncoupling of transcription with splicing in prostate cancer may explain in part the abundance of unspliced reads mapping to CE3 that we and others have reported in CSPC. These results may be result of differential interpretation when using RNA sequencing or quantitative PCR methods that do not measure across splice junctions (50). Taken together, reliable approaches for detecting AR-V7 mRNA and protein in CRPC, demonstrate that AR-V7 expression is low, and does not associate with clinical outcome, in untreated primary prostate cancer. Although multiple independent CSPC cohorts were studied, one limitation is the small number of patients across these cohorts. Importantly, assays that quantify AR-V7 in CSPC will need to be subject to intense analytical validation before they can be considered for further clinical qualification of AR-V7 as a predictive biomarker in untreated primary prostate cancer, although our data would suggest that AR-V7 testing is unlikely to have a major role to play as a predicative biomarker in this setting.

One important unanswered question, that is likely to be of therapeutic importance, is what causes AR-V7 expression in CRPC. In clinical trials of neoadjuvant-intense ADT (employing abiraterone or enzalutamide) in CSPC, these agents were not sufficient to induce significant AR-V7 expression after six months of therapy, even in patients with significant volumes of residual disease (51). We have further confirmed this in an independent cohort of patients who received neoadjuvant ADT and enzalutamide prior to radical prostatectomy, demonstrating low Ex3-CE3 *AR-V7* spliced read abundance in matched pre-and post-treatment specimens (43). In addition, *AR-V7* spliced reads did not associate with volume of residual disease following therapy, and meticulous histopathological analysis of residual disease demonstrated very rare AR-V7 staining. However, it is also important to note the challenges of low tumor cell content following endocrine therapy when performing these analyses. Furthermore, although AR-V7 protein expression is very rare and does not associate with residual tumor volume following neoadjuvant therapy, it is important to consider that it may be of biological importance in those cases where it is identified with robustly validated assays, and this warrants further investigation. In contrast to CSPC, approximately 30% of men with CRPC progressing on abiraterone or enzalutamide show detectable AR-V7 by CTC analysis, and much higher levels of AR-V7 reactivity are seen by tissue biopsy analysis following abiraterone and/or enzalutamide therapy (10, 14). Consistent with this, we show that the majority of PDX models developed from CRPC tissue biopsies demonstrate low/no AR-V7 mRNA or protein expression when grown in intact mice with a substantial increase in AR-V7 mRNA and protein abundance when castrate sublines are developed, which is reversed with testosterone treatment. These data indicate that CRPC biology supports the expression of AR-V7 mRNA and protein, but may also suggest that CSPC biology may differ with regards to induction of AR-V7 generation. Although not fully understood, these observations maybe driven by the duration of treatment (six months versus years of androgen deprivation), reactivation of AR activity to produce a rising PSA (which is not observed after six months of neoadjuvant therapy), the emergence of *AR* structural variants in CRPC such as gene and enhancer amplifications, and expression of critical cofactors (14, 51-54). A better understanding of the mechanisms that drive the emergence of AR-V7 expression in prostate cancer may provide critical insight into prostate cancer biology and support novel treatment approaches.

Overall, our study highlights the challenges of developing analytically validated and clinically qualified predictive biomarkers for prostate cancer medicine. We demonstrate that AR-V7 mRNA and protein abundance is low in CSPC prior to treatment using robustly validated assays pursuing both IHC and RNA sequencing-based approaches. Similar efforts are needed for emerging assays for detection of AR-V7 using CTCs and circulating cell-free RNA assays. Standardized controls, as well as stringent laboratory methods, are critical for both robust research discovery and determining clinical benefit. It remains to be seen whether AR-V7 can be validated and transferred to routine clinical care as a predictive biomarker in CSPC, and it maybe that the true clinical importance of AR-V7 may only be realized with the development of therapies that specifically target AR-V7 and convert it from a negative predictive to positive predictive biomarker.

## Methods

### Cell lines

22Rv1 (CRL-2505), VCaP (CRL-2876), LNCaP (CRL-1740), C42 (CRL-3314), DU145 (HTB-81) and PC3 (CRL-1435) from American Type Culture Collection, and PNT2 from Sigma-Aldrich. LNCaP95 cells were provided by Alan K. Meeker and Jun Luo (Johns Hopkins University, Baltimore, Maryland, USA). All cell lines were grown in recommended media at 37°C in 5% CO_2_. Cell lines were tested for mycoplasma using the VenorGem One Step PCR Kit (Cambio) and short tandem repeat (STR) profiled at regular intervals. Formalin-fixed, paraffin-embedded (FFPE) cell line pellets (containing 4 to 5 million cells) for immunohistochemistry were developed by fixing overnight, at 4°C, in 10% Neutral Buffered Formalin. The following day, fixed pellets were either processed through to paraffin block or transferred to 70% ethanol and kept, at 4°Celsius, until being processed.

### Immunoblotting

Briefly, cells were lysed with RIPA buffer (ThermoFischer scientific) supplemented with protease inhibitor cocktail (ThermoFischer scientific). Protein extracts (25 μg) were separated on 4/12% NuPAGE Bis-Tris gel (Invitrogen) by electrophoresis and subsequently transferred onto Immobilon-P PVDF membranes of 0.45 μm pore size (Millipore). Primary antibodies used were rabbit monoclonal anti–AR-V7 (1 in 1,000, RM7, RevMAb), rabbit monoclonal anti–AR-V7 (1 in 1,000, EPR15656, abcam), AR n-terminus (1 in 1,000, AR441, Agilent Technologies), AR c-terminus (1 in 1,000, EP6704, abcam) and GAPDH (1 in 5,000, 6C5, Santa Cruz) with species-specific secondary antibodies conjugated to horseradish peroxidase. Chemiluminescence was detected using ECL substrate (Bio-Rad) on the Chemidoc Touch imaging system (Bio-Rad).

### VCaP mouse xenograft models

VCaP mouse xenograft models have been previously described (55). Briefly, all animal studies were performed in accordance with Beth Israel Deaconess Medical Center IACUC regulations (protocol 086-2016). VCaP cells (5 million) in 100% Matrigel were injected subcutaneously into 6-week-old ICR scid mice (Taconic Biosciences). Xenografts were grown until 1,000 mm^3^; then mice were castrated. For abiraterone acetate (AA)-and enzalutamide(E)-resistant xenograft models, when castrated tumors exceeded 150% nadir volume, they were treated with AA (30 mg/kg) and E (50 mg/kg). Tumors were biopsied before castration resistance (castration-sensitive, CS), at castration resistance (castration-resistant, CR), and when resistant to AA and E therapy (AA/E resistant, AA/E R).

### Patient-derived mouse xenografts

Institute of Cancer Research patient-derived models: All animals were housed in pathogen-free facilities. All mouse work was carried out in accordance with the ICR guidelines, including approval by the ICR Animal Welfare and Ethical Review Body, and with the UK Animals (Scientific Procedures) Act 1986. The CP50 patient-derived mouse xenograft (PDX) was derived from a metastatic lymph node biopsy from a patient with CRPC who had received docetaxel, abiraterone, cabzitaxel and enzalutamide treatment for CRPC as previously described (56, 57). A second PDX, CP89, derived from a metastatic lymph node biopsy from a patient with mismatch repair deficient CRPC who had received abiraterone for CSPC and, docetaxel and enzalutamide for CRPC, was developed. A further subline of the CP50 and CP89 PDXs were developed that were grown and maintained exclusively in castrate mice (CP50C and CP89C). Experimental conditions are described in associated figure legends.

University of Washington patient-derived models: University of Washington patient-derived models: All PDX experiments were approved by the University of Washington Institutional Animal Care and Use Committee (protocol no. 3202-01). LuCaP PDX lines were established from specimens acquired at either radical prostatectomy or at autopsy, implanted, and maintained by serial passage in immune compromised male mice as described previously (42).

### Biospecimen procurement

Institute of Cancer Research and Royal Marsden patient cohorts: Patients were identified from a population of men with CRPC treated at the Royal Marsden Hospital. All patients had given written informed consent and were enrolled in institutional protocols approved by the Royal Marsden (London, UK) ethics review committee (reference no. 04/Q0801/60). Human biological samples were sourced ethically and their research use was in accordance with the terms of the informed consent provided. All tissue blocks were freshly sectioned and were only considered for IHC analyses if adequate material was present. Demographic and clinical data for each patient were retrospectively collected from the hospital electronic patient record system.

University of Washington patient cohorts: The University of Washington (UW) cohort consisted of 26 men who received radical prostatectomy without neoadjuvant therapy. A tissue microarray (TMA) of FFPE tissues with a total of 206 cores containing primary prostate acinar adenocarcinomas, tumor adjacent benign prostatic tissues and benign control tissues was generated and used in this study.

National Cancer Institute patient cohorts: The collection and analysis of tissue and demographic data from patients with high-risk localized prostate cancer treated with ADT plus enzalutamide prior to surgery was approved by the National Institutes of Health Institutional Review Board (protocol 15-c-0124). The collection and analysis of tissue and demographic data from patients with localized prostate cancer treated only by surgery was approved by the institutional review boards of Beth Israel Deaconess Medical Center (protocol 2010-P-000254/0) and the Dana-Farber/Harvard Cancer Center (protocols 15-008 and 15-492) and the University of Washington. All patients provided informed consent before participating. This research was conducted in accordance with the principles of the Declaration of Helsinki.

### Immunohistochemistry

Institute of Cancer Research and Royal Marsden immunohistochemistry (IHC): AR-V7 (RM7 RevMAb and EPR15656 abcam) and AR N-terminal (NTD) IHC assays have been previously described (14, 41). Briefly, RM7 IHC was performed using recombinant rabbit monoclonal anti-AR-V7 antibody (RM7, RevMAb). Formalin-fixed, paraffin-embedded (FFPE) cell lines, PDXs and patient tissue biopsies were first deparaffinized before antigen retrieval by microwaving (in Tris/EDTA buffer, pH 8.1) for 18 minutes at 800 W, and anti-AR-V7 antibody (1 in 500 to 1 in 50 dilution in Dako REAL diluent, Agilent Technologies) was incubated with tissue for 1 hour at room temperature. After washes, bound antibody was visualized using Dako Real EnVision Detection System (Agilent Technologies). Sections were counterstained with hematoxylin. Cell pellets from 22Rv1 (positive) and PC3 (negative) were used as controls. Rabbit IgGs were used as a further negative control. EPR15656 IHC assay was performed using recombinant rabbit monoclonal anti-AR-V7 antibody (EPR15656, abcam). Biopsies were first deparaffinized before antigen retrieval by microwaving (citrate buffer, pH6) for 18 minutes at 800 W. Blocking was performed using the protein block solution from the Novolink polymer detection system (Leica), and anti–AR-V7 antibody (1 in 200 to 1 in 50 dilution in Dako REAL diluent, Agilent Technologies) was incubated with tissue for 1 hour at room temperature. The reaction was visualised using the

Novolink polymer and DAB chromogen. Sections were counterstained with hematoxylin. Cell pellets from 22Rv1 (positive) and PC3 (negative) were used as controls. Rabbit IgGs were used as a further negative control. AR-NTD IHC was performed using mouse monoclonal anti-AR-NTD antibody (AR441, Agilent Technologies). Biopsies were first deparaffinized before antigen retrieval using pH 8.1 Tris/EDTA solution heated in a water bath, and anti-AR antibody (1 in 1,000 dilution in Dako REAL diluent, Agilent Technologies) was incubated with tissue for 1 hour at room temperature. After washes, bound antibody was visualized using Dako Real EnVision Detection System (Agilent Technologies). Sections were counterstained with hematoxylin. Cell pellets from VCaP (positive) and PC3 (negative) were used as controls. Mouse IgGs were used as a further negative control. AR C-terminal (CTD) IHC was performed using recombinant rabbit monoclonal anti-AR-CTD antibody (EP670Y, Abcam). Biopsies were first deparaffinized before antigen retrieval using a pressure cooker (Menapath Antigen Access Unit) in citrate buffer (pH 6), and anti-AR-CTD antibody (1 in 100 dilution in Dako REAL diluent, Agilent Technologies) was incubated with tissue for 1 hour at room temperature. After washes, bound antibody was visualized using Dako Real EnVision Detection System (Agilent Technologies). Sections were counterstained with hematoxylin. Cell pellets from 22Rv1 (positive) and PC3 (negative) were used as controls. Rabbit IgGs were used as a further negative control.

University of Washington immunohistochemistry: Following deparaffinization, slides were steamed for 45 min in 1x Target Retrieval Solution (Agilent technologies) and blocked with dual endogenous enzyme block for 10 min (Agilent technologies). Tissues were then incubated with anti–AR-V7 antibody (RM7 RevMAb) at 1 in 50 at 37°C for 1 hr in antibody diluent (Ventana). Primary antibody complexes were detected using the UltraVision Quanto Detection System (Thermo Fisher) as described previously (58). Tissues were counterstained with hematoxylin, mounted and imaged on a Ventana DP 200 Slide Scanner (Roche). FFPE LNCaP95 cell line pellets and LuCaP 77CR PDX tissues were used as positive controls.

National Cancer Institute immunohistochemistry: For IHC with AR [D6F11, Cell Signaling, diluted 1:200 into Biocare Renoir Red diluent (Biocare Medical)] or AR-V7 [RM7, RevMAb, diluted 1:100 into SignalStain Antibody Diluent (Cell Signaling)] IHC assay was performed in FFPE human tissue sections, slides were baked for 30 min at 60 °C, deparaffinized through xylenes and rehydrated through graded alcohols. Antigen retrieval was performed using a NxGen Decloaker (Biocare Medical), for 15 min at 110 °C Tris-EDTA Buffer, pH 9.0 (abcam) for AR-V7 or for 15 min at 110 °C in Diva Decloaker (Biocare Medical) for AR. Slides were loaded into an intelliPATH FLX autostainer (Biocare Medical). Blocking was performed with 3% hydrogen peroxide (Fisher Scientific) for 5 minutes and Background Punisher (Biocare Medical) for 10 minutes, then incubated with the primary antibody for 30 minutes at room temperature. Secondary labeling was performed with the Mach 4 Universal HRP Polymer Kit (Biocare Medical). Colorimetric detection was achieved using Betazoid DAB (Biocare Medical) for 3 minutes, and counterstaining was performed using CAT Hematoxylin (Biocare Medical). After dehydration through graded alcohols and clearing in xylenes, slides were mounted with Permount (Fisher Scientific). Cell pellets from VCaP CRPC xenografts were used as positive controls for AR-V7. Human tissue served as internal negative and positive controls for AR-NTD.

### Immunohistochemistry quantification

Nuclear and cytoplasmic AR-V7 staining was determined for PDXs and patient tissue biopsies by a pathologist (author D.N.R., B.G., M.H., or R.L.) blinded to clinical data using the modified H-score method, a semiquantitative assessment of staining intensity that reflects antigen concentration. H-score was determined according to the formula: ([% of weak staining] × 1) + ([% of moderate staining] × 2) + ([% of strong staining] × 3), yielding a range from 0 to 300 (59).

### RNA sequencing and analysis

Institute of Cancer Research and Royal Marsden RNA sequencing: CP50 and CP89 PDX experiment RNA was extracted, and RNA sequencing performed as previously described (56). Paired-end transcriptome reads were aligned to the human reference genome (GRCh38/hg38) using the STAR splice-aware aligner (v2.7.7a) with default settings for two-pass method. Reads aligning to the mouse reference genome (GRCm38/mm10) were discarded using the XenofilteR package (v1.6) (60). For AR-V7 splice junction analysis, reads mapping uniquely to Ex3-CE3 junction were quantified in the range chrX:67686127-67694672. Spliced reads per million were reported for each sample.

University of Washington RNA sequencing: Total RNA was isolated from flash frozen LuCaP PDX tissues or cell lines with RNA STAT-60 (Tel-Test) followed by purification with RNeasy Mini Kit (Qiagen) using the manufacturer’s recommended in solution DNase digestion (Qiagen). RNA concentration, purity, and integrity was assessed by NanoDrop 2000 (Thermo Fisher Scientific Inc) and 2100 Bioanalyzer (Agilent Technologies). RNA-seq libraries were constructed from 1 ug total RNA using the Illumina TruSeq Stranded mRNA LT Sample Prep Kit according to the manufacturer’s protocol. Barcoded libraries were pooled and sequenced by Illumina NovaSeq 6000 or HiSeq 2500 generating 50 bp paired end reads. Sequencing reads were mapped to the hg38 human genome and mm10 mouse genomes using the STAR.v2.7.3a 2-pass method (61). Sequences aligning to the mouse genome deriving from potential contamination with mouse tissue were removed from the analysis using XenofilteR (v1.6) (60).

National Cancer Institute RNA sequencing: FFPE tissues were lysed in 1.5 ml microcentrifuge tubes using the RNeasy FFPE Mini Kit (Qiagen) according to the manufacturer’s instructions. 100-600 ng of recovered RNA was assembled into strand-specific, paired-end, Illumina-compatible sequencing libraries using the NEBNext Ultra II Directional RNA Library Prep Kit (New England Biolabs) with the NEBNext rRNA Depletion Kit (New England Biolabs). Libraries were quantified, pooled, and sequenced paired-end on a NovaSeq S4 flowcell with 100 cycles paired-end (2×100). Demultiplexed FASTQ files were adaptor-trimmed using Trimmomatic version 0.36 (62) and aligned to GRCh38 using the STAR 2.7.0f two-pass method (61), with an average of 24.9 million mapped reads (range: 8.0 to 47.8 million mapped reads) per sample. Raw data were deposited into the NCBI Database of Genotypes and Phenotypes at https://www.ncbi.nlm.nih.gov/gap/ and can be accessed with phs001813.v2.p1 and phs001938.v3.p1.

AR splice junction analysis: Splice junction analysis was performed on cohorts from ICR/RMH, UW, and NCI, as well as cases from the GTEx, TCGA and WCDT-mCRPC cohorts. GTEx data was downloaded from the NHGRI Genomic Data Science Analysis, Visualization, and Informatics Lab-Space (AnVIL) (https://gen3.theanvil.io/) and TCGA and WCDT-mCRPC data was downloaded from the NCI Genomic Data Commons (https://gdc.cancer.gov/. Splice junctions from all cohorts were extracted from STAR-aligned BAM files using the sjFromSAMcollapseUandM.awk script in the STAR package. The precise number of reads in each alignment mapping to Ex3-Ex4 were defined by the chrX:67686127-67711401 junction, while reads mapping to Ex3-CE3 were defined by the chrX:67686127-67694672 junction. Spliced and unspliced reads mapping exclusively to CE3 were processed as described previously (49). Spliced reads or mapped reads were quantified as spliced reads per million or mapped read counts per million.

### AR-V7 signature score quantification

AR-V7 signature scores were derived using the 59-gene set reported previously (14). log2-transformed FPKM values for all human or PDX cohorts were compiled and passed to GSVA (63) for R using the parameters method=“gsva” and kcdf=“Gaussian”. The scores reported are the ES enrichment score for each sample.

### Statistical analysis

Statistical analyses were performed with GraphPad Prism version 9.2.0 (GraphPad Software). Associations between factors were measured using Spearman correlations. Comparisons of single factors between treated and untreated tumors were performed using Mann-Whitney or Welch’s t tests. Null hypothesis tests of enrichments between AR-V7 positive and negative tumors and individual dichotomous factors were performed using two-sided Fisher’s exact tests. Statistical significance was pre-specified at p < 0.05. All tests used are described in figure legends.

## Data Availability

All data produced are available from dbGaP and GEO.

https://www.ncbi.nlm.nih.gov/gap/

https://www.ncbi.nlm.nih.gov/geo/

## Acknowledgments

The authors gratefully acknowledge the patients and the families of patients who contributed to this study. This work was supported by Prostate Cancer UK (Travelling Prize Fellowship to J.M.J.V.; Research Funding to J.S.dB.), the Prostate Cancer Foundation (Young Investigator Awards to S.W., A.T.K., J.W.R., F.K., and A.Sharp; Challenge Awards to S.P.B., P.S.N., J.S.dB., A.Sharp), the Intramural Research Program of the NIH, National Cancer Institute, the Movember Foundation through the London Movember Centre of Excellence (CEO13 2-002 to J.S.dB.), the Congressionally Directed Medical Research Program Prostate Cancer Research Program (Early Investigator Research Award to J.W.R., Translational Science Award to S.R.P, and Transformative Impact Award to L.D.T), the Wellcome Trust (Clinical Research Career Development Fellowship to A.Sharp), the NIHR Biomedical Research Centre, Cancer Research UK (Centre Programme and Experimental Cancer Medicine Centre grants to J.S.dB.), the UK Department of Health, Biomedical Research Centre funding to the Royal Marsden, the Doris Duke Charitable Foundation (grant to M.C.H.), the V Foundation (to M.C.H.), the NIH (P50 CA090381 to S.P.B. and P.S.N; P50 CA090381 to S.P.B and C.T.S.; P50 CA097186 and R01 CA234715 to P.S.N.), the Veterans Affairs Research and Development Service (VA BLRD 2101BX003324 to S.R.P.), and the Lopker Family Foundation (to S.R.P.). Portions of this work utilized the computational resources of the NIH HPC Biowulf cluster. The establishment and characterization of the LuCaP patient-derived xenograft models was supported by the PNW Prostate Cancer SPORE P50CA097186, P01CA163227 and IPCR.

## Author contributions

Data acquisition: Lis, Bright, Whitlock, Trostel, Karzai, Coleman, Patel, Haffner, Russo, Figueiredo, Riisnaes, Gurel, Nava Rodrigues, Bogdan, Yuan, Plymate, Neeb, Welti, Jimenez-Vacas, Sprenger, Swain, Wilkinson, Corey, de Bono, Sharp

Analysis: Lis, Ku, Sowalsky, Coleman, Haffner, Plymate, Russo, Figueiredo, Riisnaes, Gurel, Nava Rodrigues, Bogdan, Yuan, Neeb, Jimenez-Vacas, Corey, de Bono, Sharp

Reagents/Resources: True, Nelson, Corey, Dahut

Manuscript: Sowalsky, Coleman, Patel, Haffner, Plymate, Russo, Balk, Nelson, Corey, de Bono, Sharp

Supervision: Sowalsky, Nelson, Corey, Plymate, Balk, de Bono, Sharp

