## Supplementary Figures 1-12 and Supplementary Tables 1-5 for "Assessment of Androgen Receptor splice variant-7 as a biomarker of clinical response in castration-sensitive prostate cancer"

by Adam G. Sowalsky, *et al.*

• Supplementary Tables
• Supplementary Figures

**Supplementary Tables**

**Supplementary Table 1**

| **ICR/RMH matched CSPC and CRPC cohort** |  |  |
| --- | --- | --- |
| **Baseline characteristics** |  |  |
| **CSPC biopsy** | **RevMAb**<br>**(n = 26)** | **abcam**<br>**(n = 23)** |
| **Histology (n, %)**<br>Adenocarcinoma | 26, 100% | 23, 100% |
| **Grade group (n, %)**<br>1<br>2<br>3<br>4<br>5<br>NR | 1, 3%<br>1, 3%<br>5, 19%<br>3, 12%<br>14, 54%<br>2, 8% | 1, 4%<br>1, 4%<br>4, 17%<br>3, 13%<br>12, 52%<br>2, 9% |
| **Metastatic at diagnosis (n, %)**<br>M0<br>M1<br>NR | 13, 50%<br>8, 31%<br>5, 19% | 10, 43%<br>8, 35%<br>5, 22% |
| **Treatment intent (n, %)**<br>Radical<br>Palliative | 12, 46%<br>14, 54% | 11, 48%<br>12, 52% |
| **Median diagnostic PSA (µg/l, IQR)** | 29.0, 13.3-137.0* | 28.0, 13.9-138.3* |
| **Matched CRPC biopsy** | **RevMAb**<br>**(n = 32)** | **abcam**<br>**(n = 32)** |
| **Biopsy site (n, %)**<br>Bone<br>Lymph node<br>Other | 17, 65%<br>6, 23%<br>3, 12% | 17, 65%<br>6, 23%<br>3, 12% |
| **NHT prior to biopsy (n, %)**<br>No<br>Yes | 25, 45%<br>7, 35% | 25, 45%<br>7, 35% |

**Supplementary Table 1. Clinical characteristics of the Institute of Cancer Research/Royal Marsden matched cohort of prostate cancer patients with paired castration-sensitive and castration-resistant tissue biopsies.** CSPC – castration-sensitive prostate cancer, CRPC – castration-resistant prostate cancer, n – number, NR – not recorded, AR – androgen receptor, PSA – prostate-specific antigen, IQR – interquartile range, \* – three patients without diagnostic PSA data available and one patient diagnostic PSA value > 2000 analyzed as 2000, NHT – Novel Hormonal Therapy (abiraterone and/or enzalutamide)

**Supplementary Table 2**

| **ICR/RMH primary advanced cohort** |  |
| --- | --- |
| **Baseline characteristics** |  |
| **Histology (n, %)** |  |
| Adenocarcinoma | 22, 100% |
| **Grade group (n, %)** |  |
| 1 | 0, 0% |
| 2 | 2, 9% |
| 3 | 0, 0% |
| 4 | 5, 23% |
| 5 | 15, 68% |
| **Metastatic at diagnosis (n, %)** |  |
| M0 | 2, 9% |
| M1 | 18, 82% |
| NR | 2, 9% |
| **Treatment intent (n, %)** |  |
| Radical | 0, 0% |
| Palliative | 22, 100% |
| **Median diagnostic PSA (µg/l, IQR)** | 160.5, 41.6-754.0^ |

**Supplementary Table 2. Clinical characteristics of the Institute of Cancer Research/Royal Marsden Hospital primary advanced cohort of prostate cancer patients with castration-sensitive tissue biopsies.** n – number, NR – not recorded, PSA – prostate-specific antigen, IQR – interquartile range, \* – one patient diagnostic PSA value > 2000 analyzed as 2000

**Supplementary Table 3**

| **UW primary localized cohort** |  |
| --- | --- |
| **Baseline characteristics** |  |
| **Histology (n, %)**<br>Adenocarcinoma | 26, 100% |
| **Grade group (n, %)**<br>1<br>2<br>3<br>4<br>5 | 9, 35%<br>7, 27%<br>6, 23%<br>0, 0%<br>4, 15% |
| **Tumor stage (n, %)**<br>T1<br>T2<br>T3A<br>T3B<br>T4 | 0, 0%<br>15, 58%<br>7, 27%<br>4, 15%<br>0, 0% |
| **Lymph node stage (n, %)**<br>N0<br>N1<br>Nx | 20, 77%<br>2, 8%<br>4, 15% |

**Supplementary Table 3. Clinical characteristics of the University of Washington primary localized cohort of prostate cancer patients who underwent radical prostatectomies.** n – number

**Supplementary Table 4**

| **NCI primary localized cohort** |  |
| --- | --- |
| **Baseline characteristics** |  |
| **Histology (n, %)**<br>Adenocarcinoma | 84, 100% |
| **Grade group (n, %)**<br>1<br>2<br>3<br>4<br>5 | 8, 10%<br>54, 64%<br>18, 21%<br>1, 1%<br>3, 4% |
| **Tumor stage (n, %)**<br>T1<br>T2<br>T3A<br>T3B<br>T4 | 0, 0%<br>43, 51%<br>31, 37%<br>10, 12%<br>0, 0% |
| **Lymph node stage (n, %)**<br>N0<br>N1<br>Nx | 71, 85%<br>3, 4%<br>10, 12% |
| **Median diagnostic PSA (ng/ml, IQR)** | 6.0, 5.4-8.0 |

**Supplementary Table 4. Clinical characteristics of the National Cancer Institute primary localized cohort of prostate cancer patients who underwent radical prostatectomy.** n – number, PSA – prostate-specific antigen, IQR – interquartile range

**Supplementary Table 5**

| **NCI neoadjuvant cohort** |  |
| --- | --- |
| **Baseline characteristics** |  |
| **Histology (n, %)** |  |
| Adenocarcinoma | 37, 100%* |
| **Grade group (n, %)** |  |
| 1 | 0, 0% |
| 2 | 4, 11% |
| 3 | 5, 14% |
| 4 | 11, 30% |
| 5 | 17, 46% |
| **Tumor stage (n, %)** |  |
| T1 | 4, 11% |
| T2 | 6, 16% |
| T3A | 15, 41% |
| T3B | 7, 19% |
| T4 | 5, 14% |
| **Lymph node stage (n, %)** |  |
| N0 | 26, 70% |
| N1 | 11, 30% |
| Nx | 0, 0% |
| **Median diagnostic PSA (ng/ml, IQR)** | 9.3, 5.5-17.7 |

**Supplementary Table 5. Clinical characteristics of the National Cancer Institute neoadjuvant cohort of prostate cancer patients who received neoadjuvant androgen-deprivation therapy and enzalutamide prior to radical prostatectomy.** n – number, PSA – prostate-specific antigen, IQR – interquartile range, \* - five patients had predominant adenocarcinoma with focal neuroendocrine features

#### Supplementary Figures

##### Supplementary Figure 1

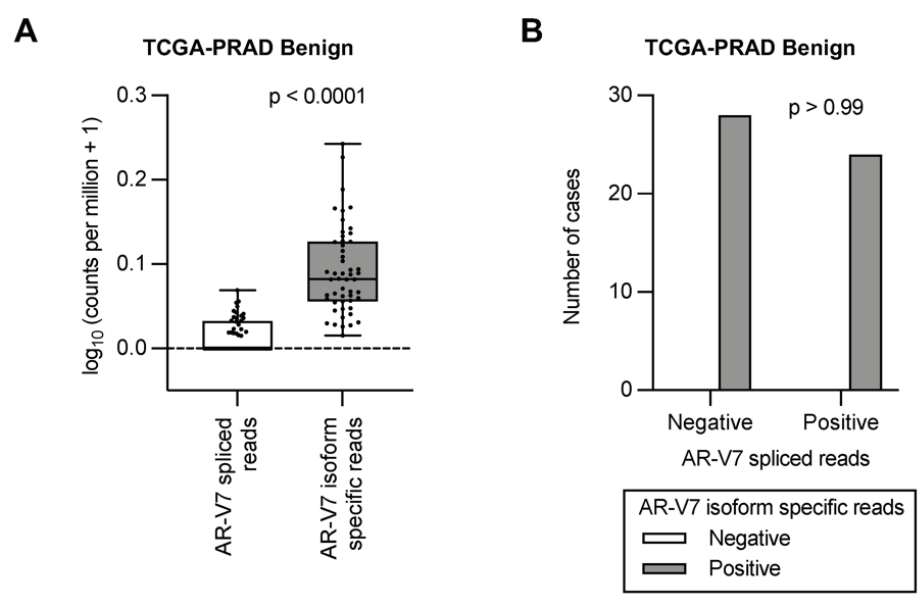

**Supplementary Figure 1. AR-V7 measurements in the benign cases of TCGA-PRAD. (A)** For each case in the TCGA-PRAD Benign ($n = 52$) the number of read counts corresponding to AR-V7 spliced reads (between exon 3 and cryptic exon 3) and AR-V7 isoform specific reads (aligning to cryptic exon 3) are shown. Spliced reads data are shown as log<sub>10</sub> (spliced reads per million + 1); isoform specific reads data are shown as log<sub>10</sub> (read counts per million + 1). Statistical significance between differences were measured by the Mann-Whitney test. Box shows median and interquartile range; bars show minimum and maximum values. **(B)** The number of cases showing concordance between the presence of AR-V7 isoform specific reads and the presence of AR-V7 spliced reads is shown. Statistical significance between associations were measured by two-sided Fisher's exact tests.

#### Supplementary Figure 2

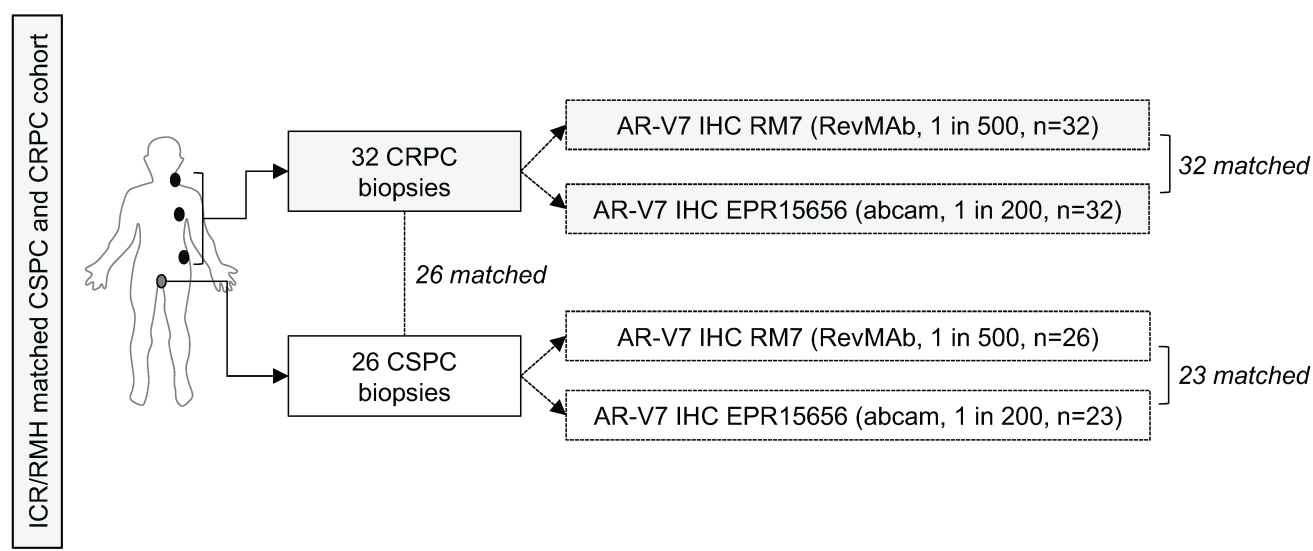

**Supplementary Figure 2. Overview of the Institute of Cancer Research/Royal Marsden Hospital (ICR/RMH) matched castration-sensitive prostate cancer (CSPC) and castration-resistant prostate cancer (CRPC) cohort.** This included 26 CSPC biopsies stained for AR-V7 (26 with RM7, RevMAb and 23 with EPR15656, abcam) and 32 CRPC biopsies (of which 26 were matched, same patient, as those CSPC biopsies utilized) stained for AR-V7 (all 32 with RM7, RevMAb and with EPR15656, abcam).

##### Supplementary Figure 3

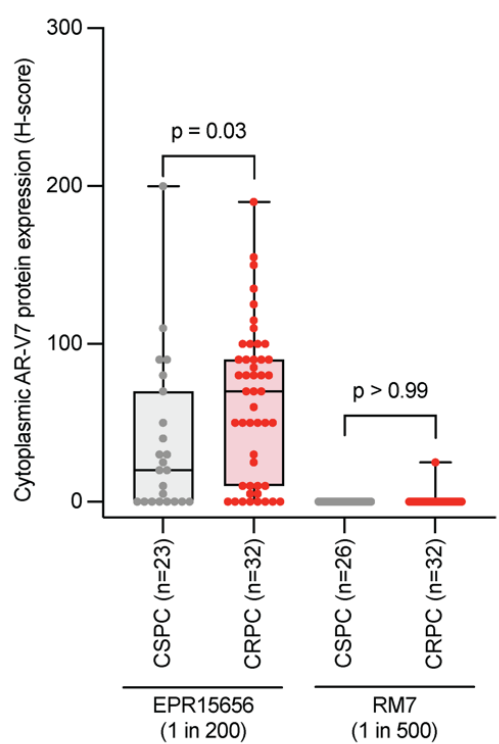

**Supplementary Figure 3. AR-V7 protein quantification by two immunohistochemistry assays in matched, same patient, castration-sensitive and castration-resistant prostate cancer tissue biopsies.** Cytoplasmic AR-V7 staining (H-score) using EPR15656 (23 CSPC and 32 CRPC) and RM7 (26 CSPC and 32 CRPC) antibodies was determined. Box shows median and interquartile range; bars show minimum and maximum values. Statistical significance between differences were measured by Mann-Whitney tests.

#### Supplementary Figure 4

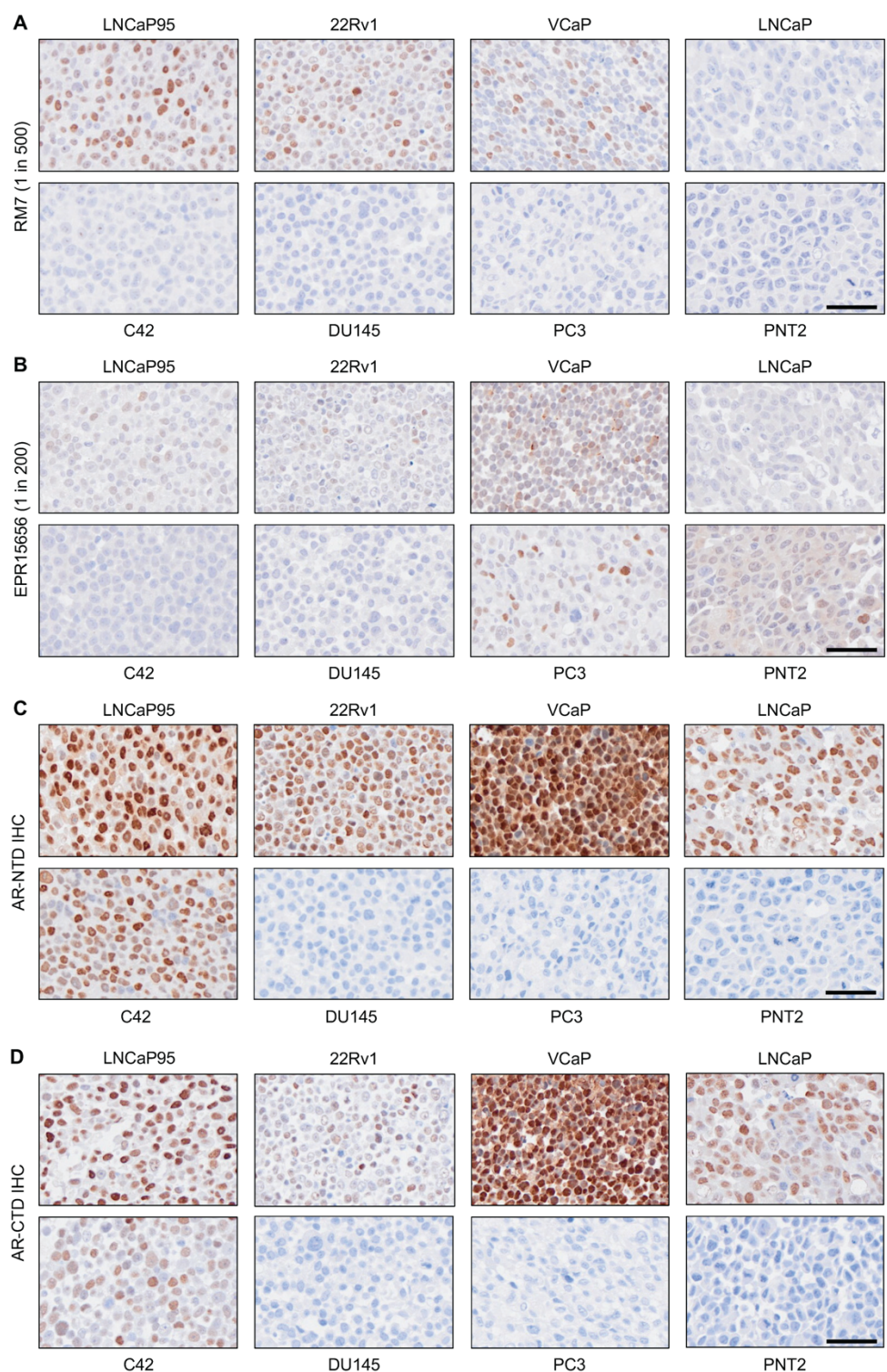

**Supplementary Figure 4. Validation of AR-V7 antibodies for AR-V7 detection by immunohistochemistry.** (A) Micrographs of AR-V7 detection by immunohistochemistry (IHC) using RM7 AR-V7 antibody (RevMAb, 1 in 500) in multiple cell line pellets (scale bar: 50 μm), (B) Micrographs of AR-V7 detection by immunohistochemistry (IHC) using EPR15656 AR-V7

#### AR-V7 in primary prostate cancer – Supplementary Information

antibody (abcam, 1 in 200) in multiple cell line pellets (scale bar: 50 $\mu\text{m}$). **(C)** Micrographs of AR N-terminal (AR-NTD) detection by immunohistochemistry (IHC) using dako AR-NTD antibody (AR441, 1 in 1000) in multiple cell line pellets (scale bar: 50 $\mu\text{m}$). **(D)** Micrographs of AR C-terminal (AR-CTD) detection by immunohistochemistry (IHC) using abcam AR-CTD antibody (EP670Y, 1 in 100) in multiple cell line pellets (scale bar: 50 $\mu\text{m}$).

#### Supplementary Figure 5

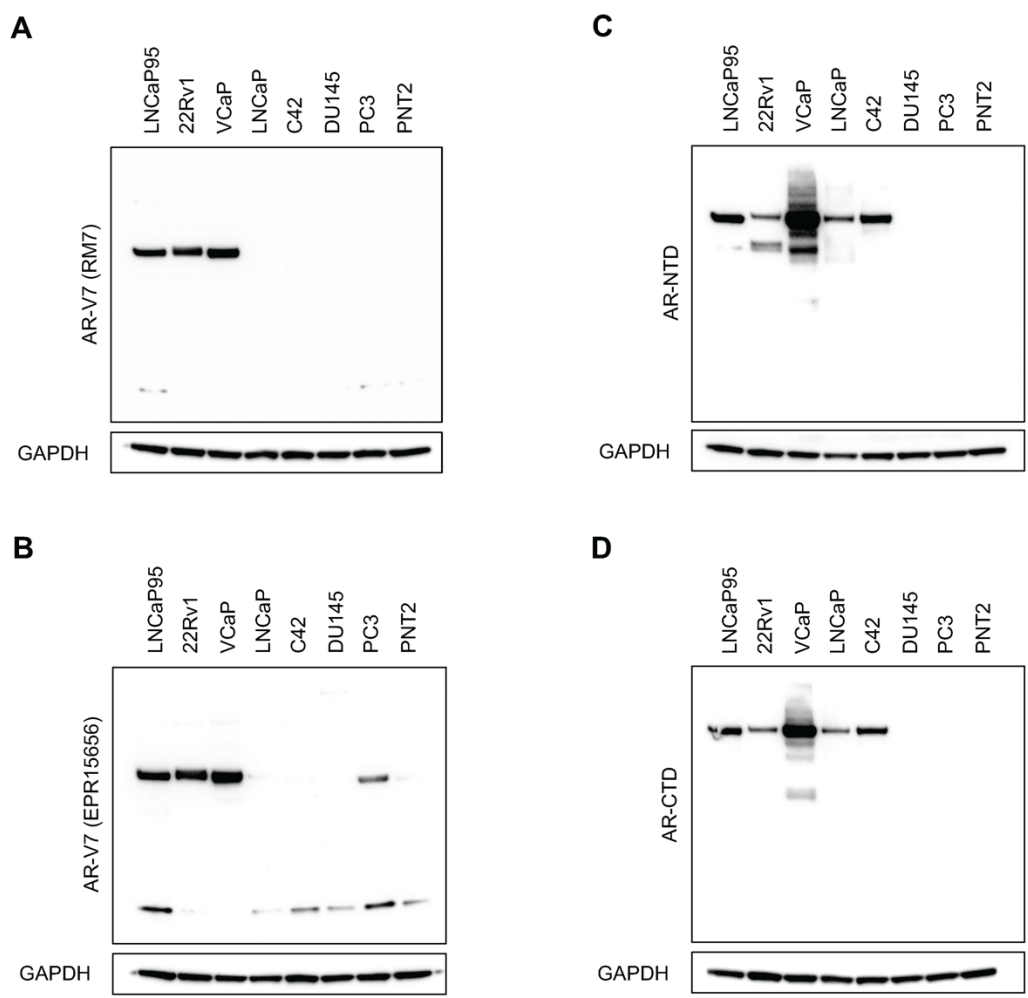

**Supplementary Figure 5. Validation of AR-V7 antibodies for AR-V7 detection by western blot.** (A) Western blot of AR-V7 detection by RM7 AR-V7 antibody (RevMAb) in multiple prostate cancer cell lines. GAPDH used as a loading control. (B) Western blot of AR-V7 detection by EPR15656 AR-V7 antibody (abcam) in multiple prostate cancer cell lines. GAPDH used as a loading control. (C) Western blot of AR-NTD detection by AR441 AR-NTD antibody (dako) in multiple prostate cancer cell lines. GAPDH used as a loading control. (D) Western blot of AR-CTD detection by EP670Y AR-CTD antibody (abcam) in multiple prostate cancer cell lines. GAPDH used as a loading control.

#### Supplementary Figure 6

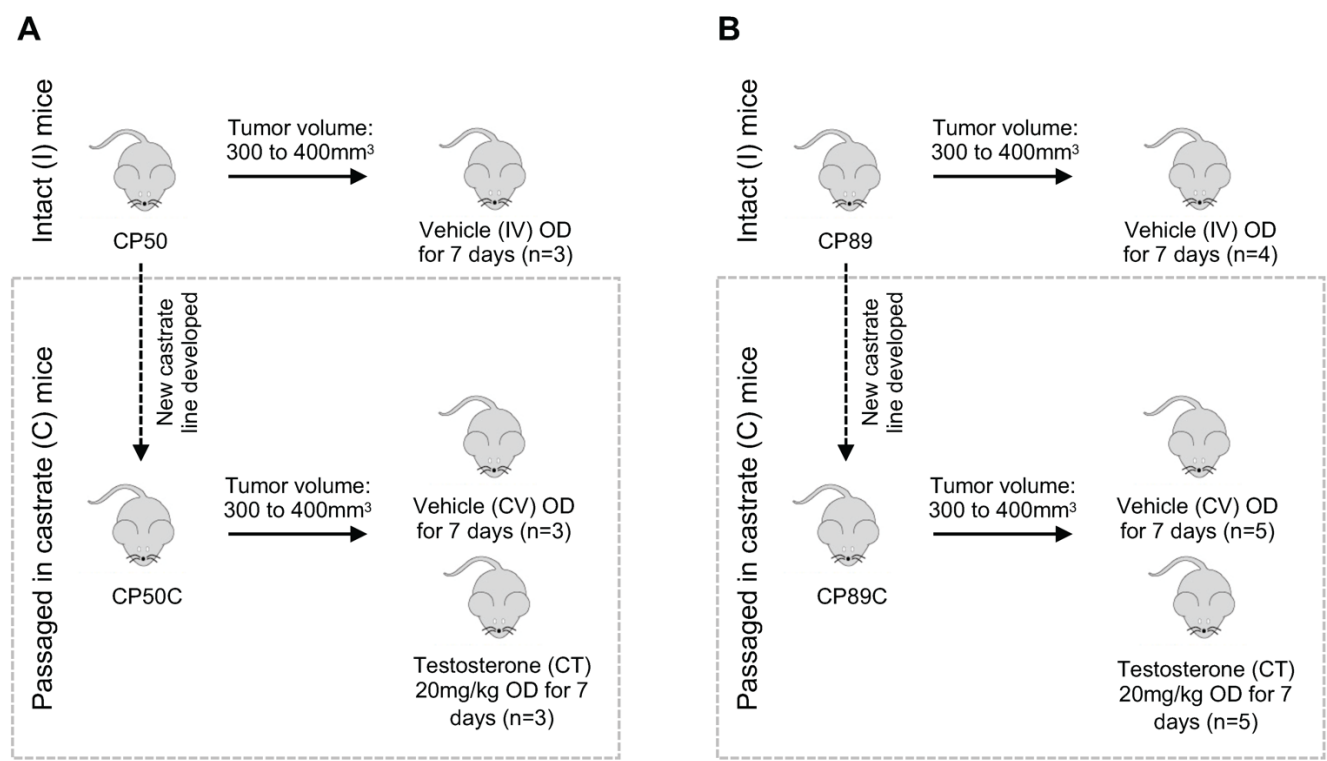

**Supplementary Figure 6. Schematic of CP50 and CP89 prostate cancer patient-derived xenograft model experimental design to determine the impact of hormonal manipulation on AR-V7 expression.** (A) CP50 patient-derived xenograft (PDX) was derived from a metastatic lymph node biopsy from a patient who had received docetaxel, abiraterone, cabazitaxel and enzalutamide for prostate cancer. CP50 was established in intact (I) mice. A further subset of CP50 PDX was developed and maintained exclusively in castrate (C) mice and termed CP50C. Once CP50 (n=3) and CP50C (n=6) PDX tumor volume reached 300 to 400mm<sup>3</sup> the CP50 PDX were treated with vehicle (intact vehicle, IV; n=3) and CP50C PDX were treated with either vehicle (castrate vehicle, CV; n=3) or 20mg/kg testosterone (castrate testosterone, CT; n=3). (B) CP89 patient-derived xenograft (PDX) was derived from a metastatic lymph node biopsy from a patient who had received abiraterone, docetaxel and enzalutamide for prostate cancer. CP89 was established in intact (I) mice. A further subset of CP89 PDX was developed and maintained exclusively in castrate (C) mice and termed CP89C. Once CP89 (n=4) and CP89C (n=10) PDX tumor volume reached 300 to 400mm<sup>3</sup> the CP50 PDX were treated with vehicle (intact vehicle, IV; n=4) and CP50C PDX were treated with either vehicle (castrate vehicle, CV; n=5) or 20mg/kg testosterone (castrate testosterone, CT; n=5).

Supplementary Figure 7

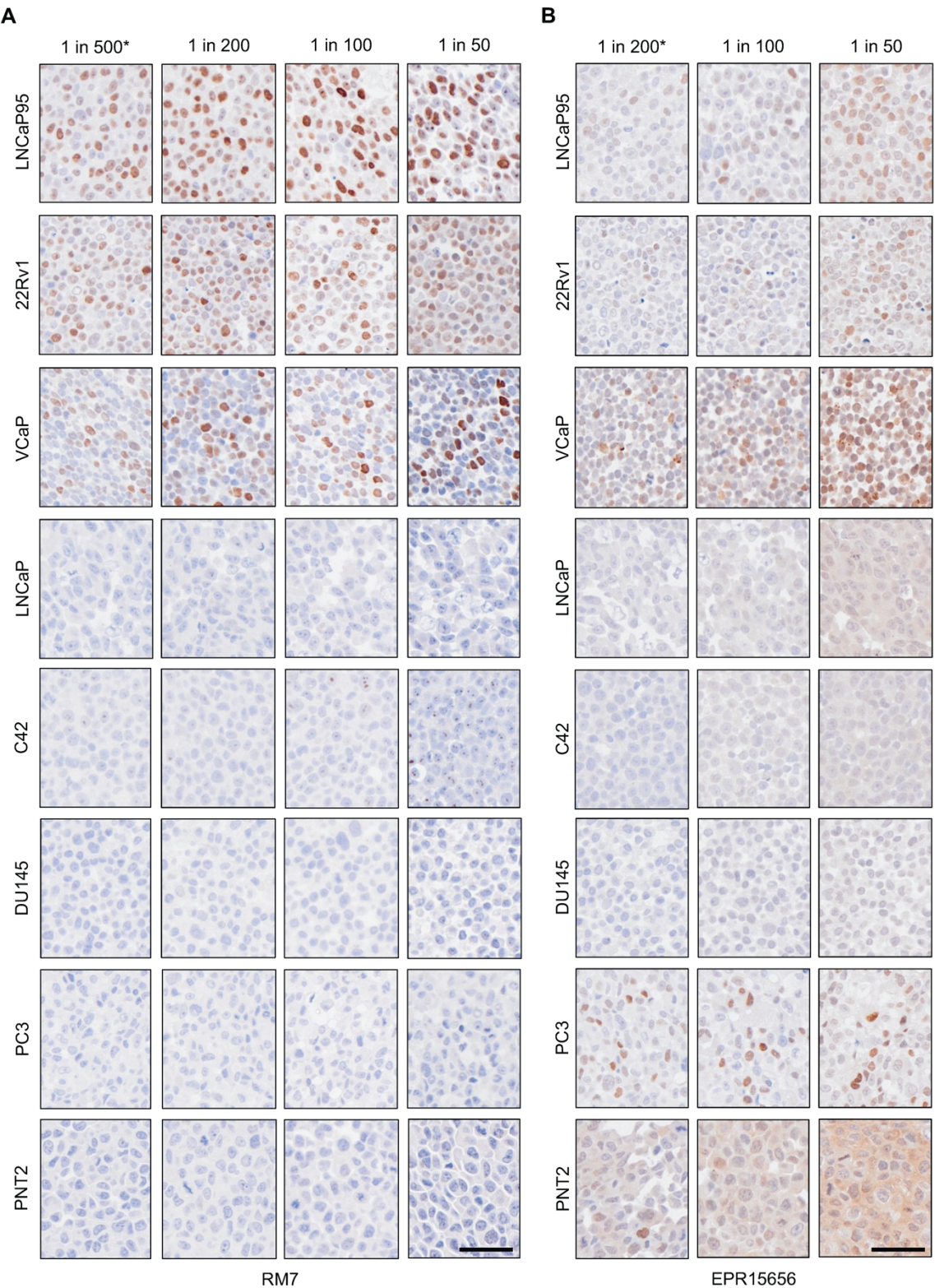

**Supplementary Figure 7. Further optimization of AR-V7 immunohistochemistry assays to enhance sensitivity and maintain specificity for detection in primary prostate cancer. (A)** Micrographs of AR-V7 detection by immunohistochemistry (IHC) using RM7 AR-V7 antibody

#### AR-V7 in primary prostate cancer – Supplementary Information

(RevMAb) at increasing concentrations (1 in 500 to 1 in 50 dilution) in multiple cell line pellets (scale bar: 50 μm). **(B)** Micrographs of AR-V7 detection by immunohistochemistry (IHC) using EPR15656 AR-V7 antibody (abcam) at increasing concentrations (1 in 500 to 1 in 50 dilution) in multiple cell line pellets (scale bar: 50 μm). \*shown in supplementary figure 4.

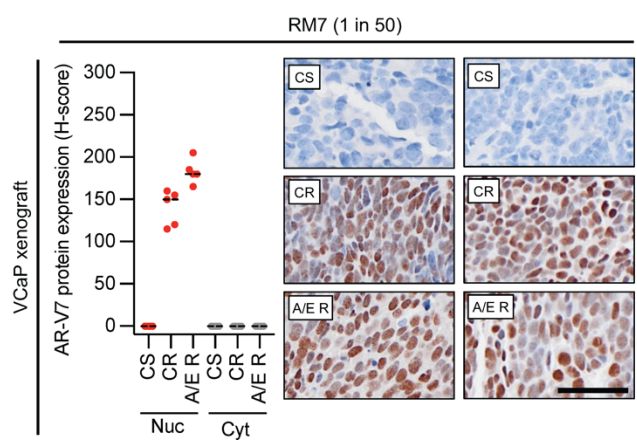

**Supplementary Figure 8. Comparison of AR-V7 immunohistochemistry assays, and AR-V7 spliced reads from RNA analysis, in the CP50 and CP89 prostate cancer patient-derived mouse xenograft, and VCaP mouse xenograft, in response to hormonal manipulation. (A)** For CP50 prostate cancer patient-derived xenograft (PDX); intact CP50 was treated with vehicle for 7 days (IV, n=3) and its castrate subline CP50C was treated with either vehicle (CV, n=3) or 20mg/kg testosterone daily (CT, n=3) for 7 days and immunohistochemistry (IHC) was performed. Representative micrographs of AR-V7 protein detection by IHC using RM7

#### AR-V7 in primary prostate cancer – Supplementary Information

(RevMAb, 1 in 50) antibody is shown (scale bar: 50 μm). Nuclear and cytoplasmic AR-V7 staining (H-score) was determined. Line represents median H-score. **(B)** For CP89 prostate cancer patient-derived xenograft (PDX); intact CP89 was treated with vehicle for 7 days (IV, n=4) and its castrate subline CP89C was treated with either vehicle (CV, n=5) or 20mg/kg testosterone daily (CT, n=5) for 7 days and IHC was performed. Representative micrographs of AR-V7 protein detection by IHC using RM7 (RevMAb, 1 in 50) antibody is shown (scale bar: 50 μm). Nuclear and cytoplasmic AR-V7 staining (H-score) was determined. Line represents median H-score. **(C)** For VCaP mouse xenografts; samples were taken from tumors that were castration-sensitive (CS, n=5), as they progressed to castration-resistant (CR, n=5), and as resistance to abiraterone and enzalutamide developed (A/E R, n=5), and IHC was performed. Representative micrographs of AR-V7 protein detection by IHC using RM7 (RevMAb, 1 in 50) antibody is shown (scale bar: 50 μm). Nuclear and cytoplasmic AR-V7 staining (H-score) was determined. Line represents median H-score.

#### Supplementary Figure 9

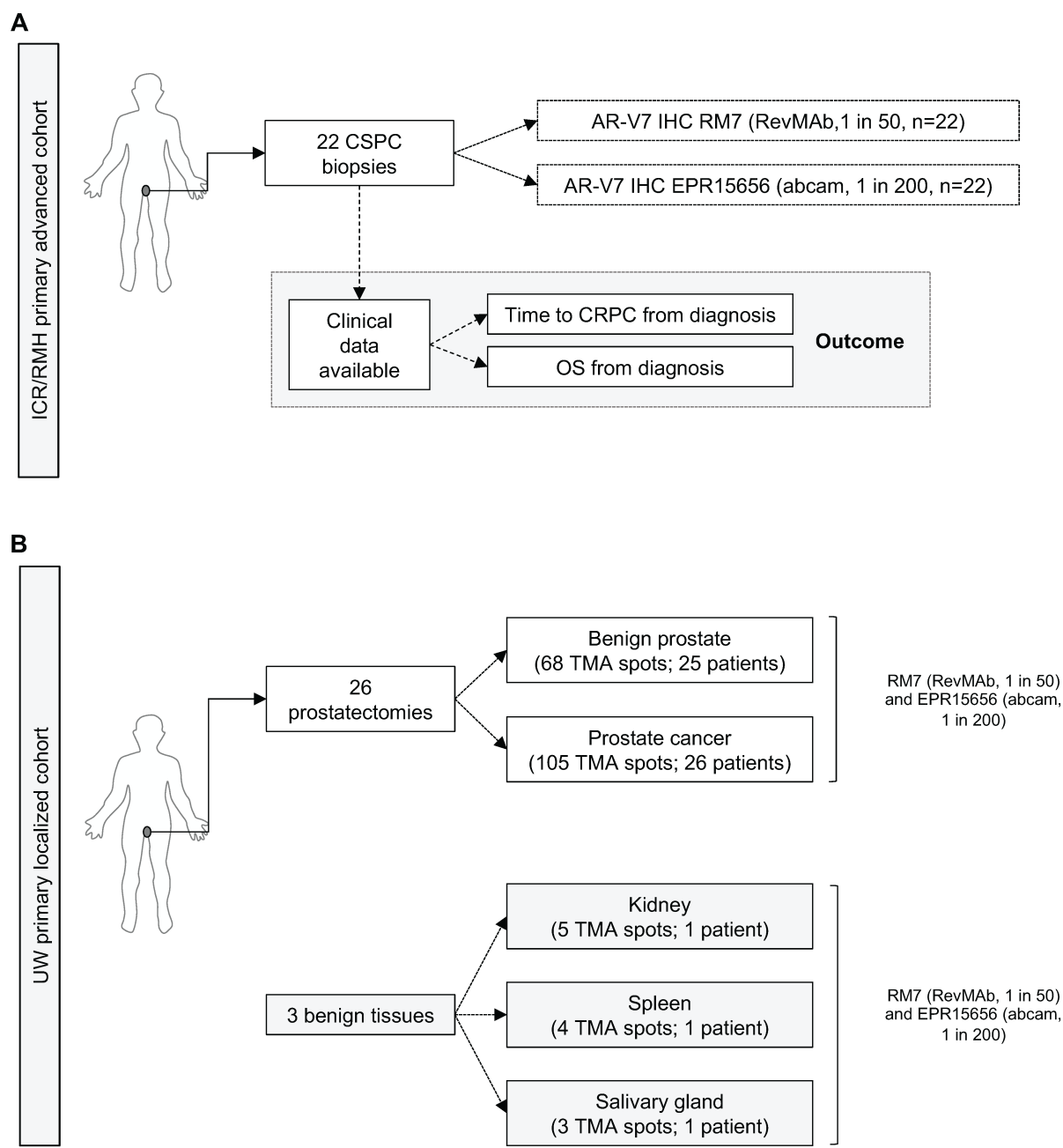

**Supplementary Figure 9. Overview of the Institute of Cancer Research/Royal Marsden primary advanced cohort and University of Washington primary localized cohort. (A)** Overview of the Institute of Cancer Research/Royal Marsden Hospital (ICR/RMH) primary advanced cohort. This included 22 diagnostic castration-sensitive prostate cancer (CSPC) biopsies stained for AR-V7 with RM7 (RevMAb, 1 in 50) and EPR15656 (abcam, 1 in 200) antibodies. All patients were treated with systemic therapy and had time to castration resistance and overall survival from diagnosis available. **(B)** Overview of the University of Washington (UW) primary localized cohort. This tissue microarray (TMA) included 26 prostatectomies with paired tumor (26 patients, 105 TMA spots) and benign (25 patients, 68 TMA spots) samples. In addition, the TMA included 3 benign tissues (kidney, 5 TMA spots; spleen, 4 TMA spots; salivary

#### AR-V7 in primary prostate cancer – Supplementary Information

gland, 3 TMA spots). The TMA was stained for AR-V7 with RM7 (RevMAb, 1 in 50) and EPR15656 (abcam, 1 in 200) antibodies.

### Supplementary Figure 10

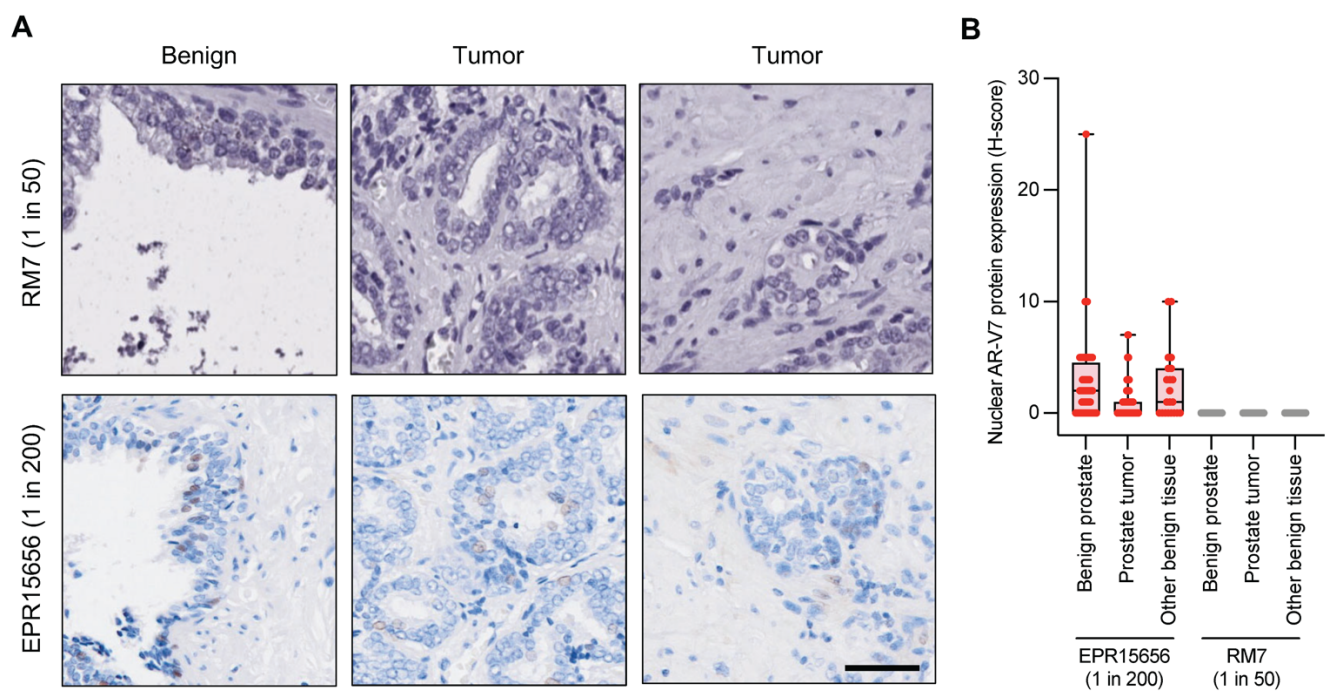

**Supplementary Figure 10. Comparison of AR-V7 protein quantification by two immunohistochemistry assays in prostate cancer patients who underwent radical prostatectomy.** (A) Representative micrographs of AR-V7 protein detection by immunohistochemistry (IHC) using EPR15656 (abcam, 1 in 200) and RM7 (RevMAb, 1 in 50) antibodies in one benign and two tumor tissue microarray (TMA) spots from radical prostatectomies from patients in the University of Washington (UW) primary localized cohort (scale bar: 50 μm). (B) Nuclear AR-V7 staining (H-score) using EPR15656 (abcam, 1 in 200, red circles) and RM7 (RevMAb, 1 in 50, gray circles) antibodies was determined. Box shows median and interquartile range; bars show minimum and maximum values.

#### Supplementary Figure 11

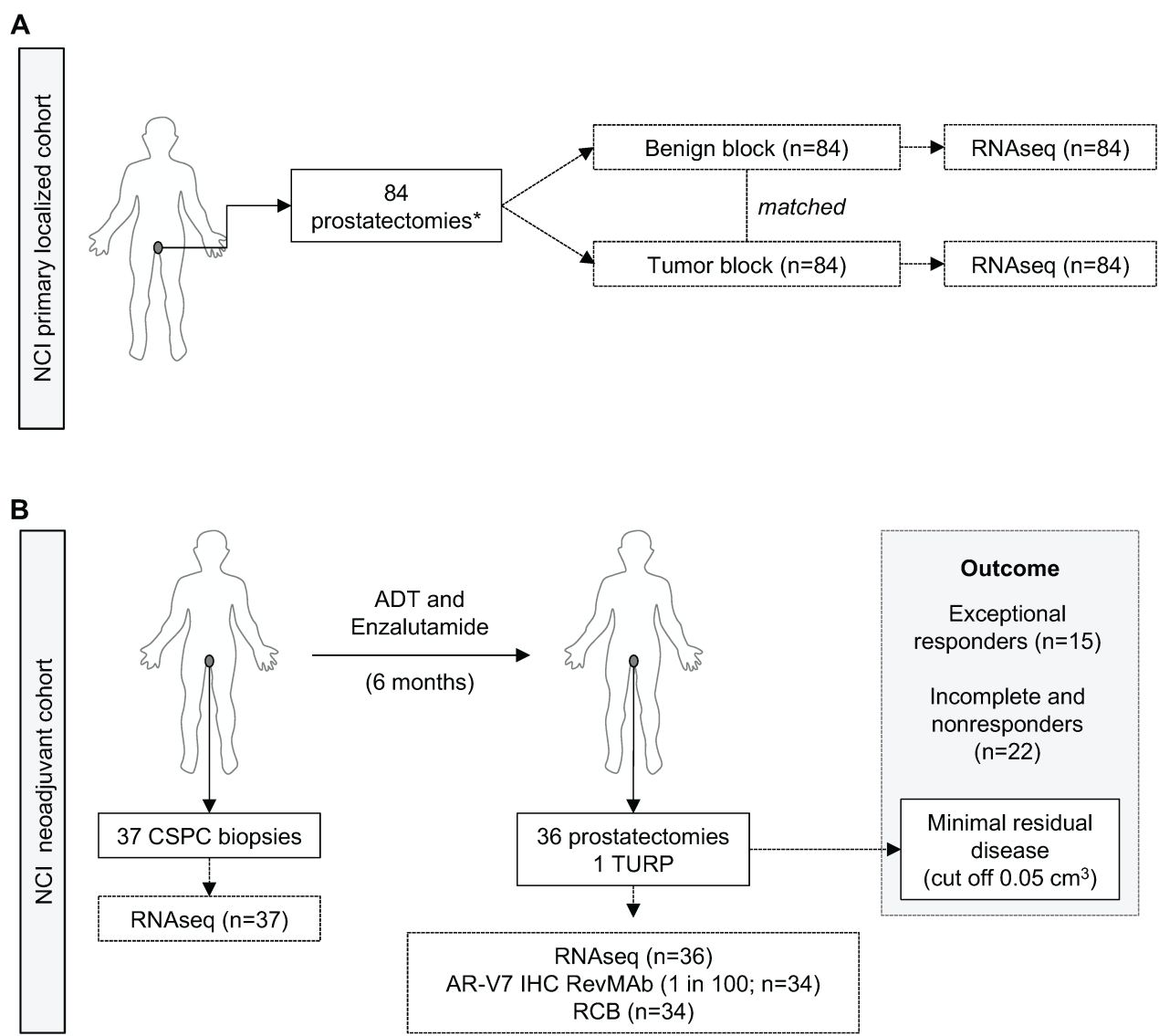

**Supplementary Figure 11. Overview of the National Cancer Institute primary localized and neoadjuvant cohorts. (A)** Overview of the National Cancer Institute (NCI) primary localized cohort. This included 84 prostatectomies with matched benign and tumor blocks with RNA sequencing analyses available. \*one patient had short course bicalutamide therapy prior to radical prostatectomy. **(B)** Overview of the NCI neoadjuvant cohort. Thirty-seven patients underwent diagnostic castration-sensitive prostate cancer (CSPC) biopsies and had RNA analyses available. Following 6 months neoadjuvant androgen-deprivation therapy (ADT) and enzalutamide 36 patients underwent radical prostatectomy and one patient underwent transurethral resection of the prostate (TURP). Of these 37 patients, by minimal residual disease (cut off 0.05cm<sup>3</sup>) on pathological examination, 15 were deemed exceptional responders and 22 were deemed incomplete or nonresponders. For these 37 patients, RNA sequencing (n=36), AR-V7 IHC by RevMAb (RM7, 1 in 100, n=34) antibody, and residual cancer burden (RCB, n=34).

#### Supplementary Figure 12

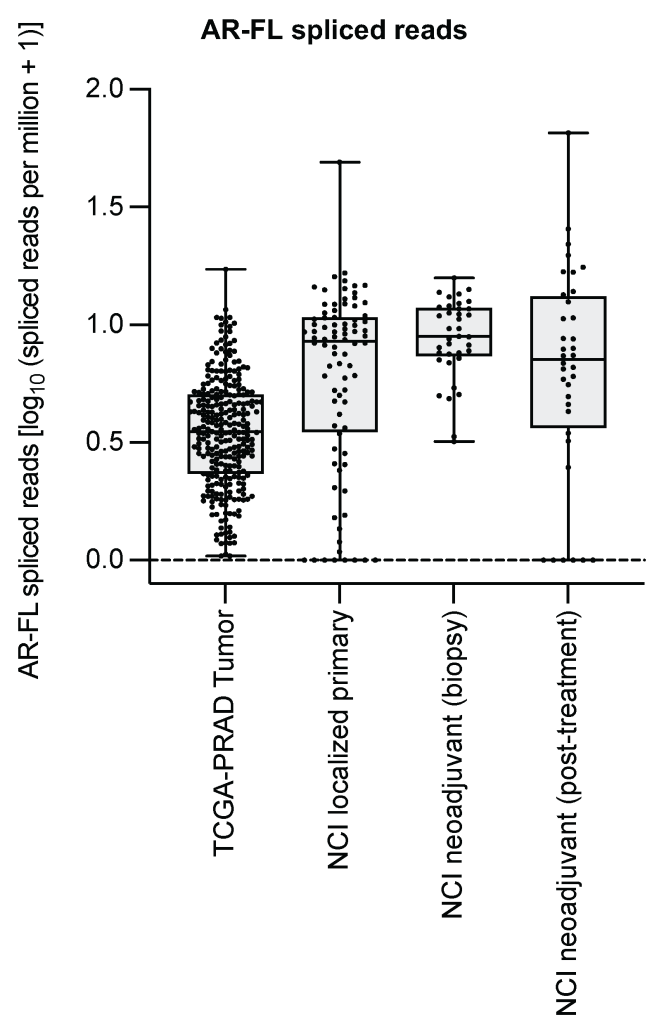

**Supplementary Figure 12. Expression of full-length AR across cohorts.** The abundance of full-length AR (AR-FL, splicing between exon 3 and exon 4) is compared across the TCGA-PRAD Tumor (n=499), National Cancer Institute (NCI) primary localized cohort (n=84), NCI neoadjuvant (biopsy, n=37), and NCI neoadjuvant (post-treatment, n=36) cohorts. Data is shown as $\log_{10} (\text{spliced reads per million} + 1)$. Box shows median and interquartile range; bars show minimum and maximum.
